# The coding of migration status in English primary care from 2011 to 2024: a pilot use of Open Code Counts

**DOI:** 10.1101/2025.07.24.25332167

**Authors:** Yamina Boukari, Lucinda Hiam, Jamie Scuffell, Arina Tamborska, Rachel Burns, Milan Wiedemann, Ines Campos-Matos, Robert W Aldridge, Sally Hargreaves, Neha Pathak, Peter Walsh, Ben Goldacre, William Hulme

## Abstract

**Background:** The migration status of the 9.8 million migrants living in England is not systematically recorded in primary care electronic health records (EHRs). Codelist approaches enable us to create cohorts of individuals who have had a predefined, optional migration-related code (e.g. “refugee”) added to their EHR.

**Aims:** We aimed to explore the use of migration-related SNOMED CT codes to inform future research using primary care data.

**Design and Setting:** We used our Open Code Counts tool and R package to explore data published by NHS England on SNOMED CT code usage in English primary care.

**Method:** We created migration-related codelists and described their use from 1st August 2011 to 31st July 2024. We compared code usage to trends in migration-related statistics from the Home Office and the 2021 Census.

**Results:** There were 29.1 million uses of 1,114 migration-related codes from 2011 to 2024. Migration-related coding increased over time, exceeding the increase observed for coding overall, with a sharp increase from 2020, particularly for country-of-birth and language.

Language coding represented 71% of code usage and where country of birth was recorded, there was mixed agreement with the census estimates. Coding of immigration legal statuses was low and overwhelmingly about asylum or refugee status.

**Conclusion:** Rapid assessment of migration-related coding using Open Code Counts highlights that a non-English first language is the most strongly represented characteristic in migrant cohorts in English primary care EHRs, which should be considered when interpreting future research findings.

**How this fits in:** This study offers population-wide insights into migration-related SNOMED CT coding in primary care in England from 2011 to 2024 using our new open-source tool, Open Code Counts. Here, we show that a first language that is not English is the most commonly recorded aspect of migration, which must be considered when interpreting results from studies that use this methodology for researching migrants’ health in primary care EHRs. We also show that migration-related coding has increased, particularly after the start of the pandemic and for country-of-birth and language codes. The increased use of these code types offers the opportunity for GP practices to better identify patients requiring language support and potential screening and service needs based on their country of birth.

**Summary sentence:** Language is the most commonly coded aspect of migration in primary care, which must be considered when interpreting primary care data studies.

## Introduction

International migrants represent 17.4% or 9.8 million individuals living in England [1]. Migration is a determinant of health, and conditions before, during and after their journey can affect health outcomes [2,3], highlighting the importance of understanding their health status and unmet health needs [4,5]. However, international migrant status (defined here as being born outside of the UK, an oft-used definition in UK-based migrant health research [6]), is not systematically or routinely recorded in primary care electronic health records (EHRs).

Several methods can be used to study migrants’ health within EHR data, such as data linkage [4,7], natural language processing [8] and codelists approaches [5,9,10]. The codelist approach involves compiling a list of predefined, optional-to-use clinical codes that relate to migration (for example, “refugee” or “interpreter needed”) and then searching the EHRs for patients who have at least one of these codes added to their record by primary care staff. Individuals with a relevant migration-related code then comprise a cohort. If migration information is instead written in free text, such as “patient is a refugee”, this individual can be missed. The results and interpretation of research based on codelists are dependent on the codes available, coding practices of individuals inputting the codes and policy initiatives that encourage or financially incentivise certain coding practices [11,12].

In previous work, a migration codelist using the Read thesaurus of clinical codes [13] was developed and validated with data from individuals registered at GP practices using the Vision EHR software (~4% of the population [14]) between 1997 and 2018 [5]. This study reported improvements in migration-related coding over time and better representation of younger migrants. Cohorts generated using this Read codelist were used to investigate allcause [9] and sexual and reproductive health-related primary care consultations and contraceptive prescriptions [15]. Another study investigated the incidence of long-term conditions in Latin American migrants using a codelist constructed from the SNOMED CT structured clinical vocabulary [16]) in a London borough [10].

Given the recent availability of population-wide primary care EHRs, such as OpenSAFELY [17], and shifts in clinical coding from Read to the more detailed SNOMED CT terminology from 2018 [16,18], it is important to investigate migration-related SNOMED CT coding at the population level. We aimed to pilot the use of a new tool developed by our team, Open Code Counts [19,20], to explore the usage of migration-related SNOMED CT codes. We then cross-referenced the findings against publicly available immigration datasets to inform the validity of future research on migrant health using codelists applied to primary care EHRs.

## Methods

### Data source: Primary care SNOMED CT coding data

We used NHS England’s annual SNOMED CT Code Usage in Primary Care datasets [21] from 1st August 2011 to 31st July 2024 within the O*pen Code Counts* R package [20] to describe the use of migration-related SNOMED CT codes. The underlying datasets represent the number of times a SNOMED CT concept was added to a GP patient record in England during the specific yearly period, with code usage rounded to the nearest 10. Calculating distinct patient counts for a given code or set of codes is not possible as a patient may have a code added to their record multiple times during the reporting period. SNOMED CT was introduced to primary care records in England in 2019, but data submitted previously in other formats (Read v2 or CTV3) were mapped to their corresponding SNOMED CT code.

### Codelists

We created lists of migration-related SNOMED CT codes using OpenCodelists.org, an openaccess website for codelist curation and sharing. To do this we implemented a previously reported search strategy to find migration-related codes [5], which were reviewed to include or exclude codes based on their relevance to the category definition (Table 1). The codelists were reviewed with two general practitioners (JS and LH). All codelists are published and openly available for reuse via OpenCodelists.org. Final codelists (comprising codes used at least once) for this analysis are included in Appendix 1.

**Table 1:** SNOMED CT codelist definitions.

| Codelist* | Definition | Search terms used in OpenCodelists.org | Number of codes included† | OpenCodelist.org reference |
| --- | --- | --- | --- | --- |
| All migration-related codes | Any code indicating that an individual could be an international migrant (e.g. related to visa or legal status, country of birth, a main language that is not English, requiring an interpreter or unable to speak/understand English or being a victim of trafficking) | abroad, asylum, born in, countr, english, exploit, forced, humanitarian, illegal, interpreter, language, leave to remain, migrant, migrat, refugee, servitude, slav, traffick, victim, visa | 2,849 | [22] |
| Country-of-birth codes | Any code indicating a country of birth that is not the UK or its devolved nations | born in | 724 | [23] |
| Immigration legal status codes | Any code indicative of a | asylum, citizenship, | 40 | [24] |
|  | specific legal status (e.g indefinite leave to remain, asylum seeker awaiting decision) | immigra, indefinite leave, refugee, spouse visa, visa, work permit |  |  |
| Asylum or refugee status codes | Any code related to refugee or asylum status, including codes related to presence in an immigration removal centre | asylum, refugee, illegal, initial health assessment | 30 | [25] |
| Language-related codes | Any code indicating a first language that is not English | language, interpreter | 1,990 | [26] |
| Interpreter need codes | Any code indicating that an individual requires an interpreter for a non-English language, (codes specifically relating to sign language were excluded) | language, interpreter | 508 | [27] |

\*Includes inactive codes and codes with 0 usage during the study period.
†Examples of individual SNOMED CT codes can be found in Table 3.

### Comparator data source: Migration statistics

We compared migration-related code usage to trends in annual migration data published by the Office for National Statistics (ONS) and the Home Office (HO; Table 2). We also compared the usage of country-of-birth codes to data on non-UK countries of birth from the Census 2021 [28].

**Table 2:** Migration datasets used for comparison against migration-related SNOMED CT coding.

| Category | Definition | Source |
| --- | --- | --- |
| Long-term international migration | Defined according to the UN definition of an international migrant as “a person who moves to a country other | ONS: Long-term international migration: Year Ending June 2012 to Year Ending June 2024, Table 1 |
|  | than that of his or her usual residence for a period of at least a year (12 months), so that the country of destination effectively becomes his or her new country of usual residence". Data represent long-term immigration to the UK for all nationalities excluding British nationality and represent annual figures ending in June of each respective year. | [29] |
| Individuals seeking asylum, on humanitarian schemes, and refugees (referred to herein as "Asylum and humanitarian status") | All individuals applying for asylum at port or in-country and individuals admitted out-of-country to a refugee resettlement scheme (including British Nationals Overseas [BNO] visas for Hong Kong Nationals and Ukraine Visa Schemes). | Asylum claims: HO table Data_Asy_D01<br>Refugee resettlement (excluding BNO and Ukraine schemes): HO table Data - Res_D02<br>BNO and Ukraine Visa Schemes: HO table Hum_01 [30] |
| Individuals granted work visas (referred to herein as "Family visa status") | All individuals issued a work-related visa out-of-country (Visa type group = "Work"; Case outcome = "Issued"). Includes main applications and dependants. | Entry clearance visa applications and outcomes detailed datasets, year ending December 2024, Data_Vis_D02 [30] |
| Individuals granted study-related visas (referred to as "Study visa status") | All individuals issued a study visa out-of-country (Visa type group = "Study"; Case outcome = "Issued"). Includes main applications and dependants. | HO: Entry clearance visa applications and outcomes detailed datasets, year ending December 2024, Data_Vis_D02 [30] |
| Individuals granted Family-related visas (referred to as "Family visa status") | All individuals issued a family visa out-of-country (Visa type group = "Work"; Case outcome = "Issued"). Includes main applications and dependants. | HO: Entry clearance visa applications and outcomes detailed datasets, year ending December 2024, Data_Vis_D02 [30] |

HO, Home Office; ONS, Office for National Statistics

### Analysis

The total numbers of recorded events for all codes that were used at least once during the study period in each codelist were plotted annually and the five most frequently used codes were tabulated. Migration-related coding as a percentage of overall SNOMED CT coding was calculated for each analysis year. The percentage increases in migration-related code usage and overall code usage during the study period were calculated as firstly, the difference in the total code usage (either migration or overall) in 2011/12 and total usage in 2023/24 over the total usage at the beginning of the period (2011/12) or secondly as annual percentage increases. Immigration data for each category of migrant were plotted and descriptively compared to the code usage data. For each of the 10 most common non-UK countries of birth according to the Census 2021, the respective percentage of code use was calculated (Census data: numerator represents the number of individuals with country of birth, denominator represents the total number of individuals with a non-UK country of birth. SNOMED CT data: numerator represents the cumulative number of the respective country’s SNOMED CT codes from 2011/12 to 2020/21; denominator represents the total recorded instances of any country-of-birth codes from 2011/12 to 2020/21).

### Tools

Data analysis was carried out using R version 4.4.2 and the *Open Code Counts* R package [20]. Analysis code was reviewed by MW and is available on GitHub [34].

## Results

### SNOMED CT coding trends

From the 1st August 2011 to 31st July 2024, there were 29.1 million (29,129,620) instances of 1,114 different migration-related codes recorded in primary care EHRs in England. Migration-related coding increased over time, with a dip at the beginning of the COVID-19 pandemic, followed by recovery and a sharp increase from 2020, particularly for country-of-birth and language-related codes, that continued until July 2024 (Figure 1). A similar increasing trend was observed for overall SNOMED CT coding (grey bar chart, Figure 1). Migration-related coding as a percentage of overall SNOMED CT coding has increased from 0.06 to 0.08% over the study period (Supplementary Figures 1-3).

**Figure 1:**
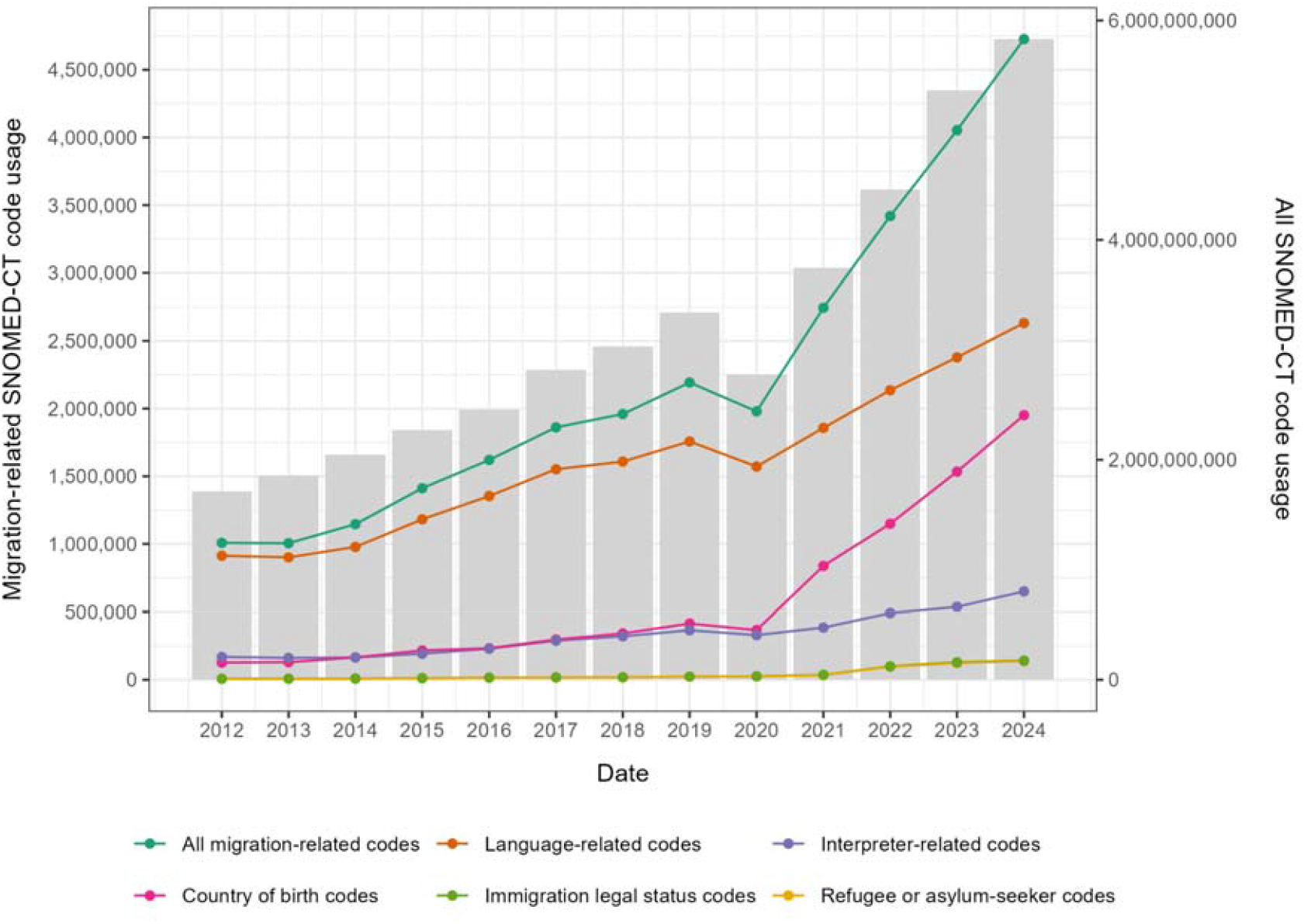
Number of migration-related SNOMED CT codes (lines) and all SNOMED CT codes (bars) recorded in primary care from 2011/12 to 2023/24.

Language codes represented the majority of migration-related codes (804/1,114 codes and 20,820,320/29,129,620 [71%] of recorded instances), with “interpreter needed” (7% of all migration-related code instances), “main spoken language Polish” (5%), “main spoken language Romanian” (4%), “main spoken language Urdu” (4%) and “interpreter present” (4%) being the most commonly used individual codes (Table 3). There were 176 codes indicating interpreter needs with 4,282,450 recorded instances.

**Table 3:** Five most commonly recorded SNOMED CT codes in each codelist.

| SNOMED CT code | Description | Number of times used | Percentage of times used within the codelist |
| --- | --- | --- | --- |
| All migration-related codes (1,114 codes with 29,129,620 recorded instances) |  |  |  |
| 315594003 | Interpreter needed (finding) | 1,913,980 | 7 |
| 315579002 | Main spoken language Polish (finding) | 1,367,000 | 5 |
| 698678003 | Main spoken language Romanian (finding) | 1,233,400 | 4 |
| 315588006 | Main spoken language Urdu (finding) | 1,083,180 | 4 |
| 314431000 | Interpreter present (finding) | 1,061,090 | 4 |
| Language-related codes (804 codes with 20,820,320 recorded instances) |  |  |  |
| 315594003 | Interpreter needed (finding) | 1,913,980 | 9 |
| 315579002 | Main spoken language Polish (finding) | 1,367,000 | 7 |
| 698678003 | Main spoken language Romanian (finding) | 1,233,400 | 6 |
| 315588006 | Main spoken language Urdu (finding) | 1,083,180 | 5 |
| 314431000 | Interpreter present (finding) | 1,061,090 | 5 |
| Interpreter codes (176 codes with 4,282,450 recorded instances) |  |  |  |
| 315594003 | Interpreter needed (finding) | 1,913,980 | 45 |
| 314431000 | Interpreter present (finding) | 1,061,090 | 25 |
| 303601000000100 | Telephone interpreting service<br>used (finding) | 432,460 | 10 |
| 736790000 | Interpreter booked<br>(finding) | 137,960 | 3 |
| 315593009 | Need for interpreter<br>(finding) | 102,370 | 2 |
| Country of birth codes (58 codes with 7,763,225 recorded instances) |  |  |  |
| 315444002 | Born in India<br>(finding) | 1,056,560 | 14 |
| 315496004 | Born in Pakistan<br>(finding) | 422,380 | 5 |
| 315491009 | Born in Nigeria<br>(finding) | 412,010 | 5 |
| 315509001 | Born in Romania<br>(finding) | 378,350 | 5 |
| 315404009 | Born in China<br>(finding) | 311,060 | 4 |
| Immigration legal status codes (23 codes with 547,730 recorded instances) |  |  |  |
| 390790000 | Asylum seeker<br>(person) | 342,490 | 63 |
| 446654005 | Refugee (person) | 107,200 | 20 |
| 728611000000100 | Asylum seeker<br>awaiting decision on<br>refugee status<br>(person) | 30,470 | 6 |
| 171420007 | Examination of<br>refugee (procedure) | 29,160 | 5 |
| 811031000000102 | Has United Kingdom<br>student visa (finding) | 9,540 | 2 |
| Asylum seeker or refugee (19 codes with 528,930 recorded instances) |  |  |  |
| 390790000 | Asylum seeker<br>(person) | 342,490 | 65 |
| 446654005 | Refugee (person) | 107,200 | 20 |
| 728611000000100 | Asylum seeker<br>awaiting decision on<br>refugee status<br>(person) | 30,470 | 6 |
| 171420007 | Examination of<br>refugee (procedure) | 29,160 | 6 |
| 748241000000103 | Unaccompanied<br>child asylum seeker<br>(person) | 9,020 | 2 |

Country-of-birth codes were the next most common code type (representing 58 unique codes and 7,763,225 recorded instances). Amongst all uses of country-of-birth codes, the five most common codes used were being born in India, Pakistan, Romania, Nigeria and China. Of the 547,730 codes relating to a specific immigration legal status, the majority (97%) indicated that an individual was an asylum seeker or refugee (Table 3); only 2% indicated that an individual had a student visa.

### Comparison of migration-related SNOMED CT coding and migration data

Long-term immigration to the UK remained relatively stable from the year ending June 2012 (565,000 individuals) to June 2021 (675,000), with a sharp increase to 1,173,000 in the year ending June 2024 (Figure 2). The number of individuals immigrating annually was numerically lower than the annual numbers of migration-related codes recorded in GP records. Of the different immigration legal status types, study (278,100 visas issues in 2012 increasing to 419,312 in 2024) and work (145,110 to 369,419) visas (for main applicants and their dependants) were the most common in the immigration data, compared to asylum-or refugee-related statuses in the coding data.

**Figure 2:**
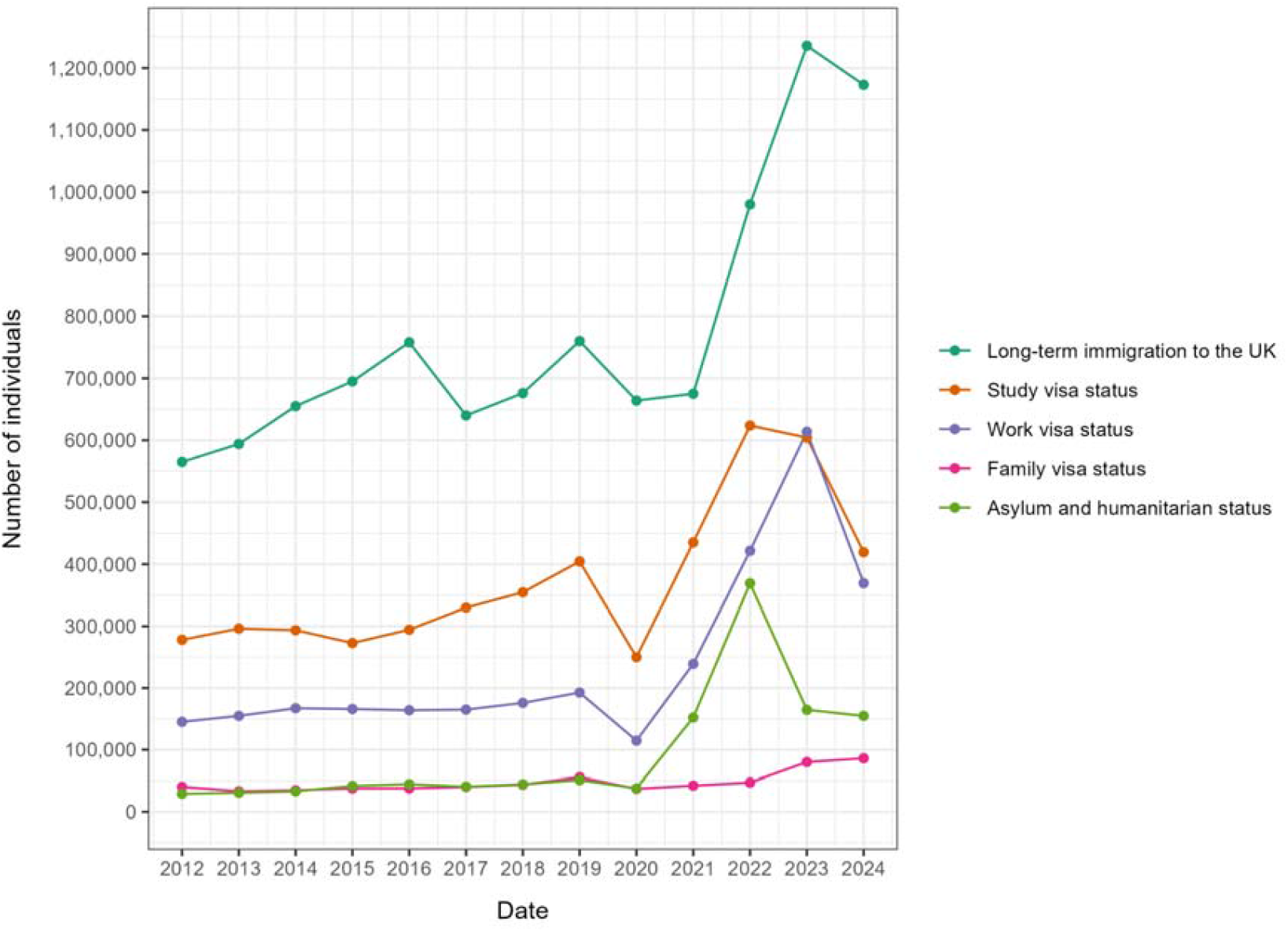
Immigration per calendar year by legal status types from 2012 to 2024. Long-term immigration [29] to the UK excludes individuals with British nationality. Study, work and family visas [30] represent grants of the respective visa types for main applicants and dependants. Asylum and humanitarian status represents asylum claims, decisions (grants, refusals and withdrawn applications), grants of out-of-country resettled refugee schemes, British Nationals Overseas [BNO] visas for Hong Kong Nationals and Ukraine Visa Schemes [30].

The percentage of “Born in India” and “Born in Nigeria” codes (as a percentage of all country-of-birth codes) used in GP data were generally consistent with the percentages of individuals born in these countries according to the 2021 Census (Figure 3). Romania, Italy and Bangladesh were overrepresented in the coding data in comparison to the Census data. Poland, Pakistan, Ireland, Germany and South Africa were underrepresented.

**Figure 3:**
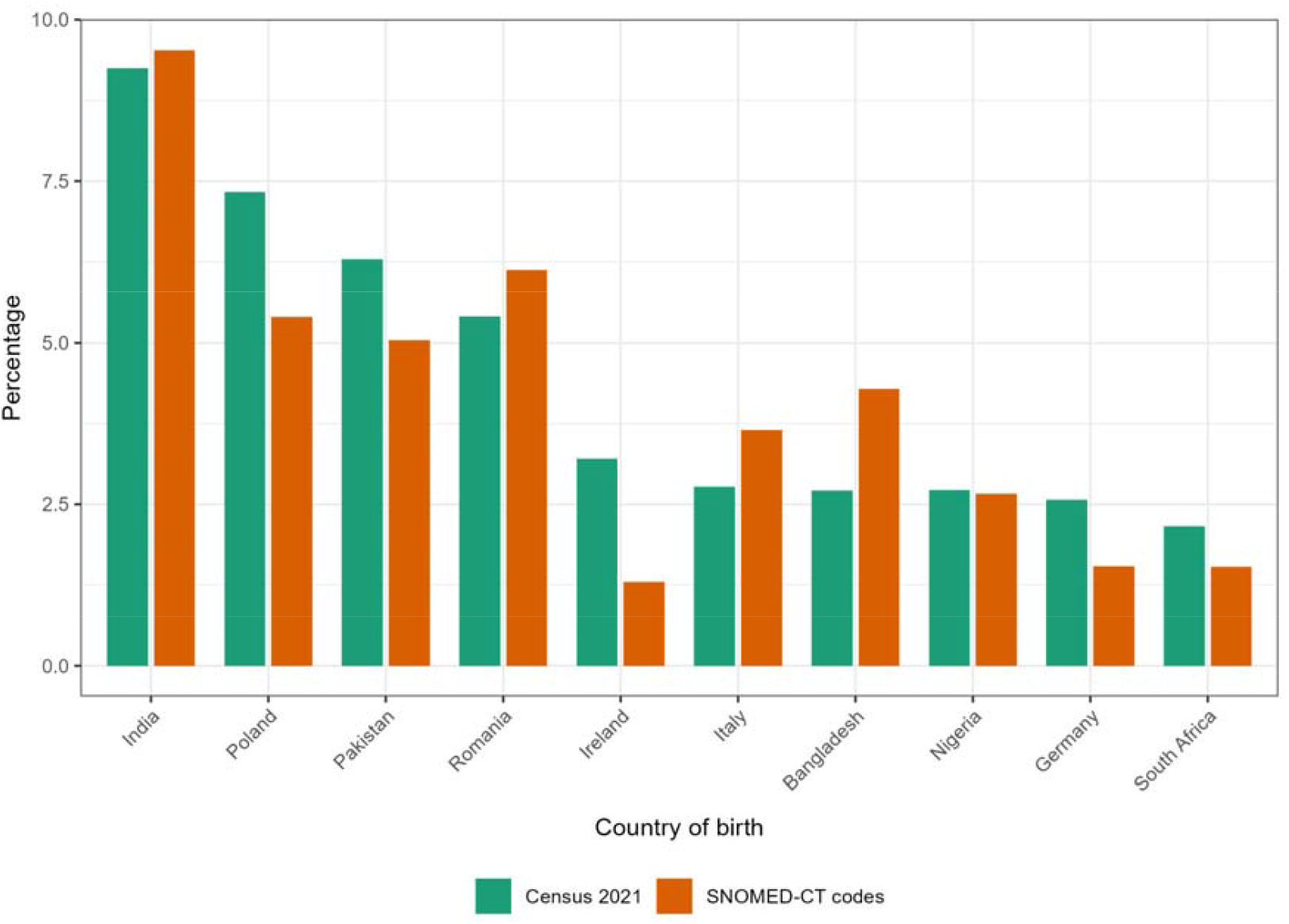
Top 10 non-UK countries of birth as a percentage of the total non-UK born population in England from the Census 2021 compared to the use of the respective country-of-birth SNOMED CT codes as a percentage of all country-of-birth SNOMED CT codes from 2011/12 to 2020/21.

## Discussion

### Summary

We found over 29 million records of migration-related codes used in primary care EHRs from 2011 to 2024 using the Open Code Counts tool. Migration-related coding has increased over time, particularly from 2020 and in the case of country-of-birth and language-related codes. Overall, language-related codes were the most commonly recorded type. Where countries of birth were recorded, there was mixed alignment with non-UK countries of birth reported in the census. Specific immigration legal statuses were less commonly recorded and when used, predominantly related to asylum or refugee status, whereas work and study statuses were more common in the HO immigration data.

### Strengths and limitations

The strength of this analysis is that it utilises data representing all uses of SNOMED CT codes across the majority of GP practices in England (n=6,624 practices covering 62,804,225 registered patients at the latest release of the data [21]). It is the first output from the Open Code Counts tool, which was designed to facilitate research in primary care data resources such as OpenSAFELY, and uses a selection of curated migration-related SNOMED CT codelists that are openly accessible for review and reuse at OpenCodelists.org.

The main limitation of this study is that data represent individual instances of code use, not individual patients (i.e. patients may have more than one migration-related code). To understand how many individuals have a migration code recorded, analysis of patient-level data is required, for example via OpenSAFELY, plans for which are underway. Additionally, we were unable to determine the drivers of migration coding for primary care staff, which is important in order to understand the potential biases in cohorts generated based on this coding, and we cannot capture free-text migration-related information, which may be where some staff chose to record migration status. Finally, this analysis represents instances of migration coding in the EHRs of individuals who have been able to register with a GP and therefore excludes individuals who may not be able to register or attend services [31,32].

### Comparison with existing literature

The gradual increase in migration coding from 2011 to 2019 is consistent with findings from the migration Read codelist [5]. At the start of the pandemic, there was a decrease in coding, consistent with reported dips in coded primary care activity at this time [33], followed by sharp increases in migration-related coding over that seen for overall coding. Potential reasons for the increase could be the push to ensure GP registration of groups such as migrants in vulnerable situations at the beginning of the pandemic, the use of EHR templates that included country of birth and main language spoken [34], a heightened awareness of migration due to the use of risk assessments related to recent travel, and the increase in all types of migration from 2021. Additionally, it may be that specific refugee resettlement schemes were provided with an initial healthcare assessment where migration-related codes were captured [35,36].

Language codes were the most commonly used migration-related code, consistent with previous findings showing that 57% of migration-related Read codes concerned language [5]. Whilst there are no formal incentives to record language, NHS England’s guidance for commissioners and the Office for Health Improvement and Disparities’ Migrant Health Guide actively encourage primary care staff to record a patient’s preferred spoken and written languages and interpreter requirements [37,38].

Compared to the Census, Romania, Italy and Bangladesh were overrepresented as countries of birth in the coding data, whereas Poland, Pakistan, Ireland, Germany and South Africa were underrepresented. The reasons for these discrepancies could reflect different health-seeking behaviours, English language skills, or coding practices of healthcare professionals. In the case of Polish and Pakistani migrants, it is possible that Polish and Urdu language codes, which were more frequently recorded than country of birth codes, were used instead.

Only 2% of all migration codes related to immigration legal statuses, which were dominated by asylum- and refugee-related statuses, contrary to immigration statistics showing that asylum and refugee statuses are one of the smaller groups of international migrants in England (29,031 asylum applications and resettled refugee status grants in 2012 and 154,690 in 2024). Potential explanations are payments for asylum seeker services being tied to recording the ‘Asylum seeker’ SNOMED CT code [36,39]. Secondly, amidst policy pushes towards inclusion health and under-served groups [40], GPs may code asylum and refugee statuses more as they consider them more relevant to the patient’s health compared to other visa statuses. Thirdly, it could reflect who primary care staff think of as migrants considering that the general public believe that 62% of all migrants come to the UK to seek asylum versus come to study, work, or join families [41].

### Implications for research and practice

In light of the prominence of language codes, migrant cohorts created using the migration codelist may be biased towards individuals who have language barriers, which should be considered when interpreting future research findings. Further qualitative and quantitative exploration of coding are needed to identify potential biases that coding practices could introduce into epidemiological findings, including the timing of migration coding and the concurrent use of different migration-related codes.

There is an opportunity to use language coding as a way to support patients, for example by prospectively identifying individuals who would benefit from double appointments to accommodate interpreter usage. Information on country of birth can help in determining screening and vaccinations requirements, and chronic disease follow-up as set out by UK migrant health guidelines [42]. The increase in country-of-birth coding from 2020 is encouraging. Qualitative work is needed to determine how to further improve country-of-birth recording, especially considering that most individuals will be asked questions regarding their country of birth and length of time in the UK when registering with a new practice but that these data are often not coded in the EHR. The use of clinical decision support systems that prompt the coding of country of birth to improve disease detection and screening guidance for migrants should be further explored [42].

Finally, financial incentives could be a way to encourage the routine recording of migration status in primary care. However, there are concerns to address regarding the historical sharing of data between NHS Digital and the Home Office for immigration enforcement purposes [43], and the impact of coding on who is charged for healthcare.

To conclude, we demonstrate that migration-related coding is nuanced and potentially captures migrants who have obvious language barriers.These nuances must be considered when interpreting research using migrant cohorts within primary care EHRs.

## Data Availability

All data referred to in this manuscript are publicly available within the Open Code Counts tool at https://bennettoxford.github.io/opencodecounts/articles/app.html

https://digital.nhs.uk/data-and-information/publications/statistical/mi-snomed-code-usage-in-primary-care

## Author contributions

Conceptualization: YB, LH. Methodology: YB, LH, JS, RB, AT, MW, WH. Software: AT, MW. Code review: MW. Formal analysis: YB. Writing - Original Draft: YB, LH. Writing - Review & Editing: YB, LH, JS, AT, RB, MW, IC-M, RWA, SH, NP, PW, BG, WH. Supervision: WH, BG. Visualization: YB. Project administration: YB. Funding acquisition: BG.

