## Supplementary Appendix for "The coding of migration status in English primary care from 2011 to 2024: a pilot use of Open Code Counts"

Supplementary Figure 1: Migration-related SNOMED-CT coding as a percentage of overall SNOMED-CT coding	2

Supplementary Figure 2: Percentage increase in migration-related and overall SNOMED-CT coding from the beginning of the study period (2011/12) to the respective year	3

Supplementary Figure 3: Annual percentage increases in migration-related and overall SNOMED-CT coding	4

Supplementary Table 1: All migration-related SNOMED CT codes used at least once between 2011-2024	5

Supplementary Table 2: Country-of-birth SNOMED CT codes used at least once between 2011-2024	46

Supplementary Table 3: Immigration legal status SNOMED CT codes used at least once between 2011-2024	56

Supplementary Table 4: Asylum or refugee status SNOMED CT codes used at least once between 2011-2024	58

Supplementary Table 5: Language-related SNOMED CT codes used at least once between 2011-2024	59

Supplementary Table 6: Interpreter need SNOMED CT codes used at least once between 2011-2024	88

### Supplementary Figure 1: Migration-related SNOMED-CT coding as a percentage of overall SNOMED-CT coding

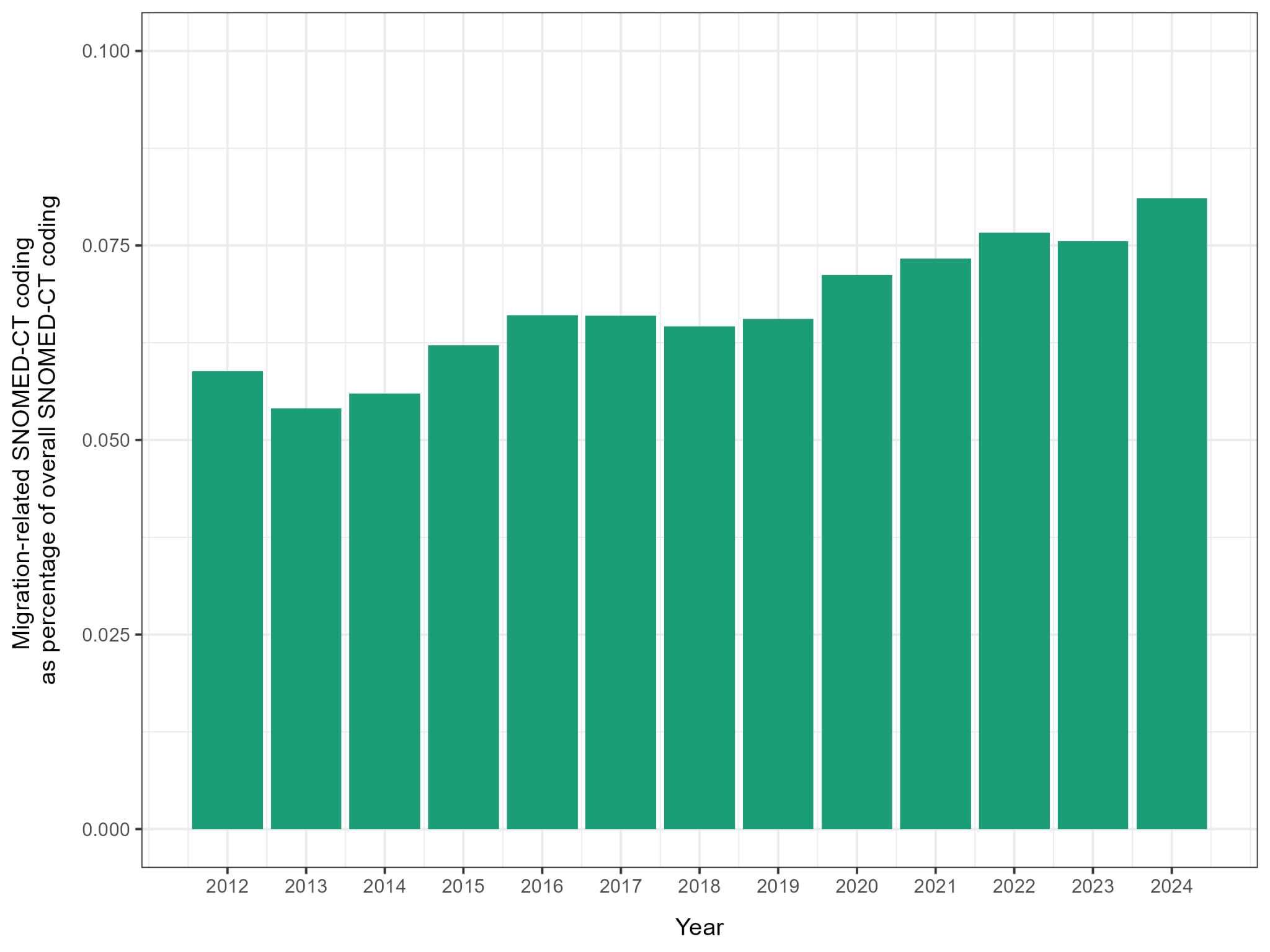

### Supplementary Figure 2: Percentage increase in migration-related and overall SNOMED-CT coding from the beginning of the study period (2011/12) to the respective year

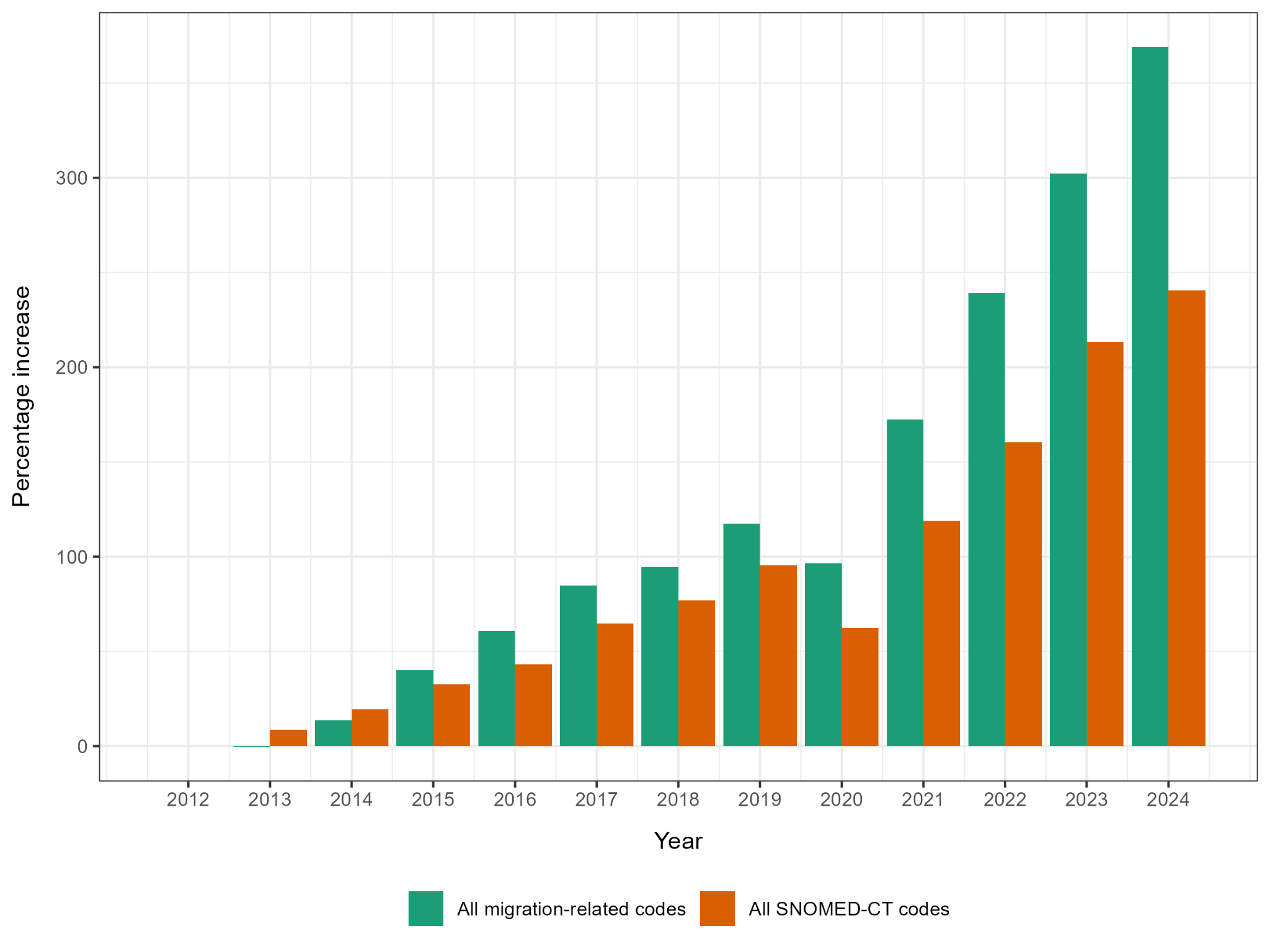

### Supplementary Figure 3: Annual percentage increases in migration-related and overall SNOMED-CT coding

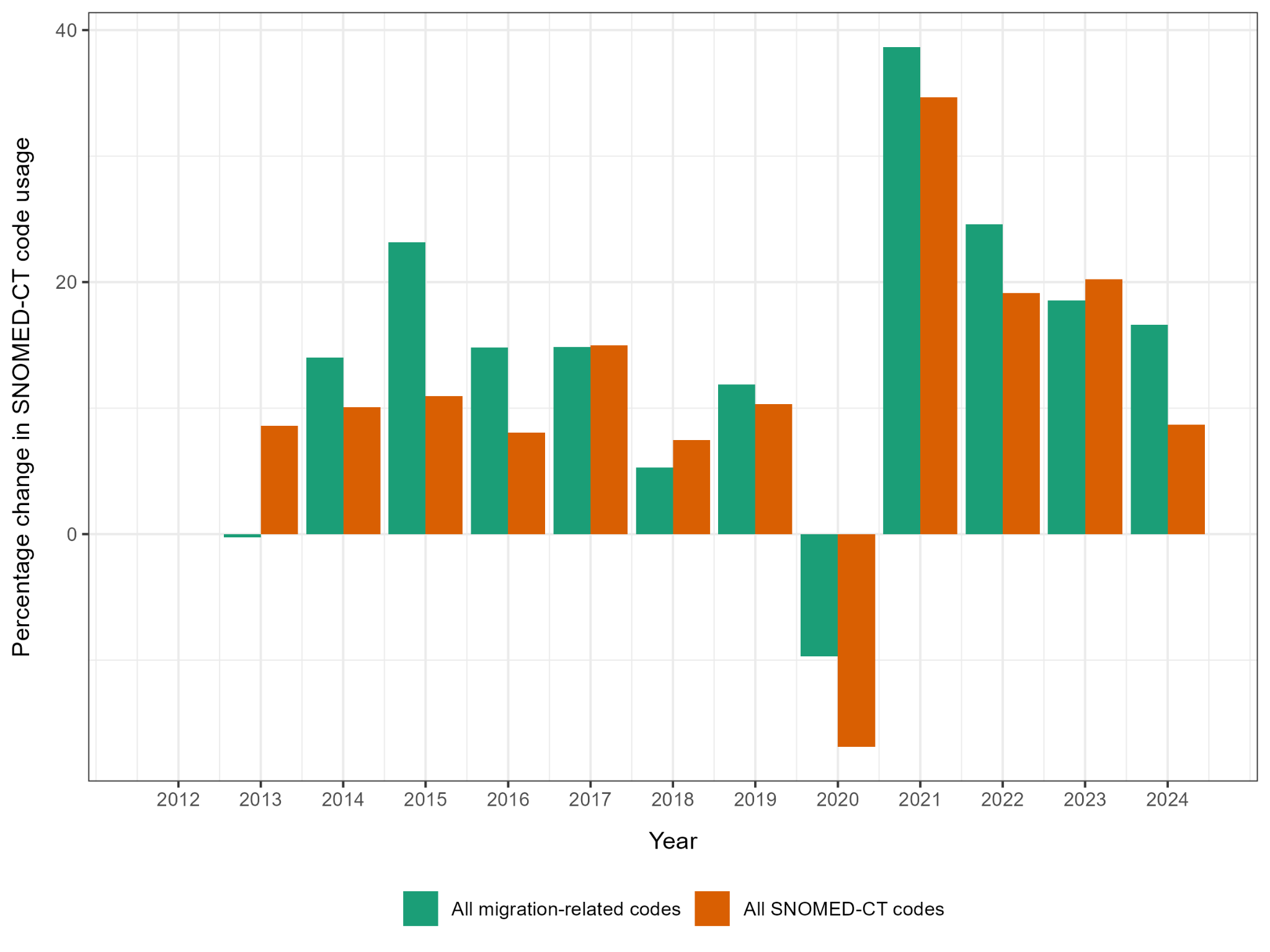

### Supplementary Table 1: All migration-related SNOMED CT codes used at least once between 2011-2024

| **SNOMED CT code** | **Description** |
| --- | --- |
| 103738006 | History and physical examination, immigration |
| 1045691000000103 | Victim of modern slavery |
| 1045861000000108 | At risk of human trafficking |
| 1045981000000106 | At risk of slavery |
| 1047281000000107 | Does not speak English |
| 1047301000000108 | Does not read English |
| 1047321000000104 | Romany language interpreter needed |
| 1057331000000104 | Signposting to Refugee Council |
| 1085811000000100 | History of detention in immigration removal centre |
| 1193634005 | Born in Cabo Verde |
| 1254713008 | Requires language interpretation service to support health literacy |
| 1364151000000105 | Immigration Removal Centre Assessment Toolkit discharge planning screening |
| 1364221000000109 | Immigration Removal Centre Assessment Toolkit reception screening |
| 1364251000000104 | Immigration Removal Centre Assessment Toolkit reception screening declined |
| 1364261000000101 | Immigration Removal Centre Assessment Toolkit secondary screening |
| 1364271000000108 | Immigration Removal Centre Assessment Toolkit secondary screening declined |
| 1364871000000107 | Immigration Removal Centre Assessment Toolkit discharge screening |
| 1364881000000109 | Immigration Removal Centre Assessment Toolkit discharge screening declined |
| 160512000 | Country: [born in] or [origin] |
| 160701002 | Social migrant |
| 160702009 | Illegal migrant |
| 161089001 | Previous countries lived in |
| 161141008 | Bengali language |
| 161142001 | Gujarati language |
| 161143006 | Hindi language |
| 161144000 | Pashtu language |
| 161145004 | Punjabi language |
| 161146003 | Urdu language |
| 161148002 | Speaks English poorly |
| 161156004 | Language difficulty |
| 161158003 | Immigrant |
| 171420007 | Examination of refugee |
| 1874641000000107 | Initial health assessment using New Patient Questionnaire for newly arrived migrants in the United Kingdom declined |
| 1874651000000105 | Initial health assessment using New Patient Questionnaire for newly arrived migrants in the United Kingdom |
| 203281000000104 | Akan language interpreter needed |
| 203291000000102 | Albanian language interpreter needed |
| 203301000000103 | Amharic language interpreter needed |
| 203311000000101 | Arabic language interpreter needed |
| 203321000000107 | Bengali language interpreter needed |
| 203371000000106 | Italian language interpreter needed |
| 203381000000108 | Cantonese language interpreter needed |
| 203391000000105 | Croatian language interpreter needed |
| 203401000000108 | Czech language interpreter needed |
| 203411000000105 | Dutch language interpreter needed |
| 203421000000104 | Persian language interpreter needed |
| 203441000000106 | French language interpreter needed |
| 203521000000103 | French Creole language interpreter needed |
| 203531000000101 | Igbo language interpreter needed |
| 203581000000102 | German language interpreter needed |
| 203591000000100 | Greek language interpreter needed |
| 203601000000106 | Gujarati language interpreter needed |
| 203611000000108 | Hakka language interpreter needed |
| 203631000000100 | Hausa language interpreter needed |
| 203641000000109 | Hebrew language interpreter needed |
| 203651000000107 | Hindi language interpreter needed |
| 203681000000101 | Japanese language interpreter needed |
| 203691000000104 | Korean language interpreter needed |
| 203701000000104 | Kurdish language interpreter needed |
| 203711000000102 | Lingala language interpreter needed |
| 203721000000108 | Lithuanian language interpreter needed |
| 203801000000105 | Ganda language interpreter needed |
| 203811000000107 | Malayalam language interpreter needed |
| 203821000000101 | Mandarin language interpreter needed |
| 203831000000104 | Norwegian language interpreter needed |
| 203841000000108 | Pashto language interpreter needed |
| 203901000000102 | Polish language interpreter needed |
| 203911000000100 | Portuguese language interpreter needed |
| 203961000000103 | Panjabi language interpreter needed |
| 203971000000105 | Russian language interpreter needed |
| 203981000000107 | Serbian language interpreter needed |
| 203991000000109 | Shona language interpreter needed |
| 204011000000102 | Sinhala language interpreter needed |
| 204021000000108 | Somali language interpreter needed |
| 204031000000105 | Spanish language interpreter needed |
| 204041000000101 | Swahili language interpreter needed |
| 204051000000103 | Swedish language interpreter needed |
| 204081000000109 | Sylheti language interpreter needed |
| 204111000000101 | Tagalog language interpreter needed |
| 204131000000109 | Tamil language interpreter needed |
| 204151000000102 | Thai language interpreter needed |
| 204171000000106 | Tigrinya language interpreter needed |
| 204191000000105 | Turkish language interpreter needed |
| 204201000000107 | Ukrainian language interpreter needed |
| 204211000000109 | Urdu language interpreter needed |
| 204221000000103 | Vietnamese language interpreter needed |
| 204241000000105 | Yoruba language interpreter needed |
| 205091000000107 | Born in Eritrea |
| 224619008 | Migrant |
| 224628009 | Hinko language |
| 297289008 | World languages |
| 297290004 | Afro-Asiatic language |
| 297291000 | Berber language |
| 297292007 | Kabyle language |
| 297296005 | Tamazight language |
| 297298006 | Chadic language |
| 297299003 | Hausa language |
| 297300006 | Cushitic language |
| 297301005 | Afar language |
| 297303008 | Oromo language |
| 297304002 | Saho language |
| 297305001 | Sidamo language |
| 297306000 | Somali language |
| 297308004 | Egyptian language |
| 297309007 | Coptic language |
| 297310002 | Semitic language |
| 297311003 | Aramaic language |
| 297312005 | Assyrian language |
| 297313000 | Syriac language |
| 297314006 | Canaanitic language |
| 297315007 | Hebrew language |
| 297316008 | Ethiopic language |
| 297317004 | Amharic language |
| 297319001 | Gurage language |
| 297320007 | Harari language |
| 297321006 | Tigre language |
| 297322004 | Tigrinya language |
| 297323009 | North Arabic language |
| 297324003 | Arabic language |
| 297325002 | Classical Arabic language |
| 297326001 | South Arabic language |
| 297327005 | Maltese language |
| 297328000 | Altaic language |
| 297329008 | Mongolian language |
| 297332006 | Tungusic language |
| 297337000 | Manchu language |
| 297339002 | Sibo language |
| 297340000 | Turkic language |
| 297341001 | Chuvash language |
| 297344009 | Khakass language |
| 297345005 | Tuvinian language |
| 297347002 | Balkar language |
| 297349004 | Kara-Kalpak language |
| 297350004 | Karachai language |
| 297351000 | Kazakh language |
| 297352007 | Kirgiz language |
| 297353002 | Kumyk language |
| 297355009 | Tatar language |
| 297358006 | Salar language |
| 297359003 | Uigur language |
| 297360008 | Uzbek language |
| 297362000 | Azerbaijani language |
| 297363005 | Turkish language |
| 297364004 | Turkmen language |
| 297367006 | Esperanto language |
| 297368001 | Interlingua language |
| 297370005 | Australian language |
| 297371009 | Aranda language |
| 297373007 | Caucasian language |
| 297375000 | Avar language |
| 297376004 | Dargin language |
| 297378003 | Lezgin language |
| 297380009 | Eastern Caucasian language |
| 297381008 | Chechen language |
| 297382001 | Ingush language |
| 297383006 | Southern Caucasian language |
| 297384000 | Georgian language |
| 297385004 | Western Caucasian language |
| 297389005 | Circassian language |
| 297391002 | Central and South American Indian language |
| 297392009 | Andean equatorial language |
| 297395006 | Aymara language |
| 297396007 | Guarani language |
| 297398008 | Quechua language |
| 297400007 | Ge-Pano-Carib language |
| 297401006 | Carib language |
| 297405002 | Tacana language |
| 297407005 | Bribri language |
| 297408000 | Cabecar language |
| 297411004 | Lenca language |
| 297414007 | Central Sudanic language |
| 297415008 | Lugbara language |
| 297416009 | Madi language |
| 297417000 | Mangbetu language |
| 297418005 | Sara language |
| 297419002 | Eastern Sudanic language |
| 297420008 | Nilotic language |
| 297421007 | Eastern Nilotic language |
| 297422000 | Bari language |
| 297424004 | Lotuko language |
| 297425003 | Masai language |
| 297426002 | Teso language |
| 297429009 | Kalenjin language |
| 297430004 | Suk language |
| 297431000 | Western Nilotic language |
| 297432007 | Acholi language |
| 297434008 | Dinka language |
| 297435009 | Lango language |
| 297436005 | Luo language |
| 297437001 | Nuer language |
| 297439003 | Nubian language |
| 297440001 | Djerma language |
| 297441002 | Dravidian language |
| 297442009 | Brahui language |
| 297444005 | Kanarese language |
| 297446007 | Kurukh language |
| 297447003 | Malayalam language |
| 297448008 | Tamil language |
| 297449000 | Telugu language |
| 297450000 | Tulu language |
| 297454009 | Fur language |
| 297455005 | Independent language |
| 297457002 | Barushaski language |
| 297459004 | Basque language |
| 297460009 | Japanese language |
| 297461008 | Korean language |
| 297463006 | Vietnamese language |
| 297464000 | Indo-European language |
| 297465004 | Albanian language |
| 297466003 | Armenian language |
| 297467007 | Baltic language |
| 297468002 | Latvian language |
| 297469005 | Lithuanian language |
| 297475001 | Irish Gaelic language |
| 297477009 | Germanic language |
| 297478004 | Northern Germanic language |
| 297479007 | Danish language |
| 297480005 | Faroese language |
| 297481009 | Icelandic language |
| 297482002 | Norwegian language |
| 297483007 | Swedish language |
| 297484001 | Western Germanic language |
| 297485000 | Afrikaans language |
| 297486004 | Dutch language |
| 297488003 | Flemish language |
| 297490002 | German language |
| 297491003 | Luxembourgian language |
| 297492005 | Yiddish language |
| 297493000 | Hellenic language |
| 297494006 | Greek language |
| 297495007 | Italic language |
| 297496008 | Latin language |
| 297497004 | Romance language |
| 297498009 | Catalan language |
| 297499001 | French language |
| 297501009 | Swiss French dialect |
| 297502002 | Italian language |
| 297503007 | Moldavian language |
| 297504001 | Portuguese language |
| 297505000 | Galician Portuguese dialect |
| 297507008 | Rhaeto-Romanic language |
| 297508003 | Rumanian language |
| 297509006 | Sardinian language |
| 297510001 | Spanish language |
| 297511002 | Slavic language |
| 297512009 | Eastern Slavic language |
| 297513004 | Belorussian language |
| 297514005 | Russian language |
| 297515006 | Ukrainian language |
| 297516007 | Southern Slavic language |
| 297517003 | Bulgarian language |
| 297518008 | Macedonian language |
| 297519000 | Serbo-Croatian language |
| 297520006 | Slovenian language |
| 297521005 | Western Slavic language |
| 297522003 | Czech language |
| 297523008 | Polish language |
| 297524002 | Slovak language |
| 297525001 | Sorbian language |
| 297526000 | Indo-Iranian language |
| 297527009 | Indic language |
| 297528004 | Assamese language |
| 297530002 | Sylhety |
| 297531003 | Bihari language |
| 297532005 | Hindustani language |
| 297533000 | Kashmiri language |
| 297534006 | Konkani language |
| 297535007 | Maldivian language |
| 297536008 | Marathi language |
| 297537004 | Nepali language |
| 297538009 | Oriya language |
| 297539001 | Pakistani punjabi language |
| 297540004 | Lehnda punjabi language |
| 297541000 | Sikh punjabi language |
| 297542007 | Rajasthani language |
| 297543002 | Romany language |
| 297544008 | Sanskrit language |
| 297545009 | Sindhi language |
| 297546005 | Sinhalese language |
| 297547001 | Iranian language |
| 297548006 | Baluchi language |
| 297549003 | Gilaki language |
| 297550003 | Kurdish language |
| 297551004 | Mazanderani language |
| 297552006 | Ossetian language |
| 297553001 | Persian language |
| 297554007 | Tajik language |
| 297555008 | Khoisan language |
| 297556009 | Bushman language |
| 297557000 | Hatsa language |
| 297561006 | Maban language |
| 297562004 | Maba language |
| 297563009 | Malayo-Polynesian language |
| 297564003 | Indonesian language |
| 297565002 | Achinese language |
| 297566001 | Balinese language |
| 297568000 | Bikol language |
| 297569008 | Buginese language |
| 297570009 | Cham language |
| 297571008 | Chamorro language |
| 297573006 | Igorot language |
| 297575004 | Javanese language |
| 297576003 | Madurese language |
| 297577007 | Malagasy language |
| 297578002 | Malay language |
| 297580008 | Minangkabau language |
| 297582000 | Pampangan language |
| 297583005 | Pangasinan language |
| 297584004 | Rhade language |
| 297585003 | Sundanese language |
| 297586002 | Tagalog language |
| 297587006 | Visayan language |
| 297592008 | Ponapean language |
| 297593003 | Trukese language |
| 297595005 | Melanesian language |
| 297596006 | Fijian language |
| 297598007 | Tolai language |
| 297600001 | Polynesian language |
| 297601002 | Hawaiian language |
| 297602009 | Māori language |
| 297603004 | Marquesan language |
| 297604005 | Niuean language |
| 297606007 | Samoan language |
| 297607003 | Tahitian language |
| 297608008 | Tongan language |
| 297612002 | Bahnar language |
| 297613007 | Khasi language |
| 297614001 | Khmer language |
| 297615000 | Mon language |
| 297620000 | Munda language |
| 297624009 | Santali language |
| 297625005 | Savara language |
| 297626006 | Niger-Congo language |
| 297627002 | Adamawa-Eastern language |
| 297628007 | Adamawa language |
| 297630009 | Eastern language (Niger-Congo) |
| 297631008 | Banda language |
| 297632001 | Gbaya language |
| 297633006 | Sango language |
| 297634000 | Zande language |
| 297635004 | Benue-Congo language |
| 297637007 | Bemba language |
| 297639005 | Bulu language |
| 297640007 | Chagga language |
| 297641006 | Chiga language |
| 297643009 | Duala language |
| 297645002 | Ganda language |
| 297646001 | Gisu language |
| 297648000 | Herero language |
| 297649008 | Kamba language |
| 297650008 | Kikuyu language |
| 297651007 | Kisii language |
| 297652000 | Kongo language |
| 297653005 | Lingala language |
| 297655003 | Lozi language |
| 297657006 | Luhya language |
| 297658001 | Lunda language |
| 297664008 | Meru language |
| 297666005 | Ndebele language |
| 297667001 | Ngala language |
| 297670002 | Nyanja language |
| 297672005 | Pedi language |
| 297673000 | Ruanda language |
| 297674006 | Rundi language |
| 297675007 | Shona language |
| 297676008 | Sotho language |
| 297678009 | Swahili language |
| 297679001 | Swazi language |
| 297681004 | Toro language |
| 297682006 | Tsonga language |
| 297683001 | Tswana language |
| 297684007 | Tumbuka language |
| 297685008 | Venda language |
| 297686009 | Xhosa language |
| 297687000 | Yao language - Bantu |
| 297689002 | Zulu language |
| 297690006 | Efik language |
| 297691005 | Ibibio language |
| 297692003 | Tiv language |
| 297693008 | Ijo language |
| 297694002 | Western Sudanic language |
| 297695001 | Gur language |
| 297696000 | Bariba language |
| 297697009 | Dagomba language |
| 297698004 | Gurma language |
| 297702000 | Kwa language |
| 297703005 | Adangme language |
| 297705003 | Bassa language |
| 297707006 | Edo language |
| 297708001 | Ewe language |
| 297710004 | Fanti language |
| 297711000 | Fon language |
| 297712007 | Ga language |
| 297714008 | Grebo language |
| 297715009 | Ibo language |
| 297716005 | Idoma language |
| 297717001 | Kru language |
| 297718006 | Nupe language |
| 297719003 | Twi language |
| 297720009 | Urhobo language |
| 297721008 | Yoruba language |
| 297722001 | Mande language |
| 297723006 | Bambara language |
| 297726003 | Loma language |
| 297727007 | Malinke language |
| 297728002 | Mende language |
| 297729005 | Soninke language |
| 297730000 | Susu language |
| 297733003 | Balante language |
| 297735005 | Fulani language |
| 297736006 | Gola language |
| 297737002 | Kissi language |
| 297738007 | Serer language |
| 297739004 | Temne language |
| 297740002 | Wolof language |
| 297741003 | Native North American language |
| 297742005 | Algonkian language |
| 297745007 | Cheyenne language |
| 297746008 | Cree language |
| 297747004 | Delaware language |
| 297748009 | Fox language |
| 297752009 | Ojibwa language |
| 297753004 | Passamaquoddy language |
| 297757003 | Chilcotin language |
| 297758008 | Chipewyan language |
| 297769006 | Seneca language |
| 297771006 | Mayan language |
| 297777005 | Mam language |
| 297778000 | Maya language |
| 297782003 | Muskogean language |
| 297783008 | Chickasaw language |
| 297785001 | Creek language |
| 297787009 | Oto-Manguean language |
| 297788004 | Chinantec language |
| 297792006 | Otomi language |
| 297794007 | Penutian language |
| 297796009 | Nez Perce language |
| 297800003 | Flathead language |
| 297801004 | Lillooet language |
| 297808005 | Omaha language |
| 297809002 | Osage language |
| 297812004 | Tanoan language |
| 297813009 | Tewa language |
| 297814003 | Tiwa language |
| 297823000 | Mayo language |
| 297824006 | Nahuatl language |
| 297832003 | Kwakiutl language |
| 297838004 | Mixe language |
| 297841008 | Zunian language |
| 297844000 | Chukchi language |
| 297849005 | Yukagir language |
| 297850005 | Papuan language |
| 297853007 | Enga language |
| 297854001 | Hagen language |
| 297855000 | Kate language |
| 297856004 | Marind language |
| 297858003 | Pidgin and Créole language |
| 297860001 | French Créole language |
| 297862009 | Krio language |
| 297863004 | Papiamento language |
| 297864005 | Pidgin English language |
| 297865006 | Police Motu language |
| 297867003 | Saramacca language |
| 297869000 | Saharan language |
| 297870004 | Kanuri language |
| 297872007 | Sino-Tibetan language |
| 297873002 | Miao-Yao language |
| 297875009 | Sinitic language |
| 297876005 | Chinese language |
| 297877001 | Cantonese Chinese dialect |
| 297878006 | Fukienese dialect |
| 297881001 | Hakka dialect |
| 297883003 | Mandarin dialect |
| 297885005 | Tai language |
| 297887002 | Lao language |
| 297889004 | Puyi language |
| 297890008 | Shan language |
| 297891007 | Thai language |
| 297893005 | Tibeto-Burman language |
| 297894004 | Bodo language |
| 297895003 | Burmese language |
| 297896002 | Chin language |
| 297897006 | Garo language |
| 297899009 | Kachin language |
| 297900004 | Karen language |
| 297901000 | Lahu language |
| 297902007 | Lepcha language |
| 297904008 | Lushei language |
| 297905009 | Meithei language |
| 297907001 | Nakhi language |
| 297908006 | Newari language |
| 297909003 | Tibetan language |
| 297910008 | Yi language |
| 297911007 | Songhai language |
| 297912000 | Uralic language |
| 297914004 | Finnic language |
| 297915003 | Estonian language |
| 297916002 | Finnish language |
| 297920003 | Mordvin language |
| 297923001 | Hungarian language |
| 297927000 | Enets language |
| 297928005 | Nenets language |
| 297931006 | Language commonly spoken in Europe |
| 297934003 | Swiss German dialect |
| 298018001 | Kashubian Polish dialect |
| 303601000000100 | Telephone interpreting service used |
| 306211000000109 | Failed asylum seeker |
| 313421002 | Créole language |
| 313422009 | Mirpuri language |
| 314430004 | Presence of interpreter |
| 314431000 | Interpreter present |
| 315355003 | Country of birth - European |
| 315356002 | Country of birth - Asian |
| 315357006 | Country of birth - American continent |
| 315358001 | Country of birth - African |
| 315359009 | Country of birth - Australasian |
| 315360004 | Country of birth - Atlantic |
| 315361000 | Country of birth - Pacific |
| 315365009 | Born in Afghanistan |
| 315366005 | Born in Albania |
| 315367001 | Born in Algeria |
| 315368006 | Born in Andorra |
| 315369003 | Born in Angola |
| 315370002 | Born in Antigua and Barbuda |
| 315372005 | Born in Argentina |
| 315373000 | Born in Armenia |
| 315374006 | Born in Australia |
| 315375007 | Born in Austria |
| 315376008 | Born in Azerbaijan |
| 315377004 | Born in Bahamas |
| 315378009 | Born in Bahrain |
| 315379001 | Born in Bangladesh |
| 315380003 | Born in Barbados |
| 315381004 | Born in Belgium |
| 315382006 | Born in Belize |
| 315383001 | Born in Belorussia |
| 315384007 | Born in Benin |
| 315385008 | Born in Bhutan |
| 315386009 | Born in Bolivia |
| 315387000 | Born in Bosnia - Herzegovnia |
| 315388005 | Born in Botswana |
| 315389002 | Born in Brazil |
| 315390006 | Born in British Guyana |
| 315391005 | Born in Brunei |
| 315392003 | Born in Bulgaria |
| 315393008 | Born in Burkina Faso |
| 315394002 | Born in Burma |
| 315395001 | Born in Burundi |
| 315396000 | Born in Cambodia |
| 315397009 | Born in Cameroon |
| 315398004 | Born in Canada |
| 315399007 | Born in Cape Verde Islands |
| 315400000 | Born in Central African Republic |
| 315401001 | Born in Chad |
| 315402008 | Born in Chechnya |
| 315403003 | Born in Chile |
| 315404009 | Born in China |
| 315405005 | Born in Colombia |
| 315406006 | Born in Comoros Islands |
| 315407002 | Born in Congo |
| 315408007 | Born in Costa Rica |
| 315409004 | Born in Croatia |
| 315410009 | Born in Cuba |
| 315411008 | Born in Cyprus |
| 315412001 | Born in Czech Republic |
| 315413006 | Born in Democratic People's Republic of Korea |
| 315414000 | Born in Denmark |
| 315415004 | Born in Djibouti |
| 315416003 | Born in Dominican Republic |
| 315417007 | Born in East Timor |
| 315418002 | Born in Ecuador |
| 315419005 | Born in Egypt |
| 315420004 | Born in El Salvador |
| 315422007 | Born in Equatorial Guinea |
| 315423002 | Born in Estonia |
| 315424008 | Born in Ethiopia |
| 315425009 | Born in Fiji |
| 315426005 | Born in Finland |
| 315427001 | Born in France |
| 315428006 | Born in Gabon |
| 315429003 | Born in Gambia |
| 315430008 | Born in Georgia |
| 315431007 | Born in Germany |
| 315432000 | Born in Ghana |
| 315433005 | Born in Greece |
| 315434004 | Born in Grenada |
| 315435003 | Born in Guatemala |
| 315436002 | Born in Guinea Bissau |
| 315437006 | Born in Guinea Republic |
| 315438001 | Born in Guyana |
| 315439009 | Born in Haiti |
| 315440006 | Born in Honduras |
| 315441005 | Born in Hong Kong |
| 315442003 | Born in Hungary |
| 315443008 | Born in Iceland |
| 315444002 | Born in India |
| 315445001 | Born in Indonesia |
| 315446000 | Born in Iran |
| 315447009 | Born in Iraq |
| 315448004 | Born in Ireland |
| 315449007 | Born in Israel |
| 315450007 | Born in Italy |
| 315451006 | Born in Ivory Coast |
| 315452004 | Born in Jamaica |
| 315453009 | Born in Japan |
| 315454003 | Born in Jordan |
| 315455002 | Born in Kazakhstan |
| 315456001 | Born in Kenya |
| 315457005 | Born in Kiribati |
| 315458000 | Born in Kosovo |
| 315459008 | Born in Kuwait |
| 315460003 | Born in Kyrgyzstan |
| 315461004 | Born in Laos |
| 315462006 | Born in Latvia |
| 315463001 | Born in Lebanon |
| 315464007 | Born in Lesotho |
| 315465008 | Born in Liberia |
| 315466009 | Born in Libya |
| 315467000 | Born in Liechtenstein |
| 315468005 | Born in Lithuania |
| 315469002 | Born in Luxembourg |
| 315470001 | Born in Madagascar |
| 315471002 | Born in Malawi |
| 315473004 | Born in Malaysia |
| 315474005 | Born in Maldives |
| 315475006 | Born in Mali |
| 315476007 | Born in Malta |
| 315477003 | Born in Mauritania |
| 315478008 | Born in Mauritius |
| 315479000 | Born in Mexico |
| 315480002 | Born in Moldavia |
| 315481003 | Born in Monaco |
| 315482005 | Born in Mongolia |
| 315483000 | Born in Morocco |
| 315484006 | Born in Mozambique |
| 315485007 | Born in Namibia |
| 315486008 | Born in Nauru |
| 315487004 | Born in Nepal |
| 315488009 | Born in New Zealand |
| 315489001 | Born in Nicaragua |
| 315490005 | Born in Niger |
| 315491009 | Born in Nigeria |
| 315492002 | Born in North Korea |
| 315494001 | Born in Norway |
| 315495000 | Born in Oman |
| 315496004 | Born in Pakistan |
| 315497008 | Born in Palestine |
| 315498003 | Born in Panama |
| 315499006 | Born in Papua New Guinea |
| 315500002 | Born in Paraguay |
| 315501003 | Born in Peru |
| 315502005 | Born in Philippines |
| 315503000 | Born in Poland |
| 315504006 | Born in Portugal |
| 315505007 | Born in Puerto Rico |
| 315506008 | Born in Qatar |
| 315507004 | Born in Republic of Ireland |
| 315508009 | Born in Republic of Korea |
| 315509001 | Born in Romania |
| 315510006 | Born in Russia |
| 315511005 | Born in Rwanda |
| 315512003 | Born in San Marino |
| 315513008 | Born in Sao Tome and Principe |
| 315514002 | Born in Saudi Arabia |
| 315516000 | Born in Senegal |
| 315517009 | Born in Seychelles |
| 315518004 | Born in Sierra Leone |
| 315519007 | Born in Singapore |
| 315520001 | Born in Slovakia |
| 315521002 | Born in Slovenia |
| 315522009 | Born in Solomon Islands |
| 315523004 | Born in Somalia |
| 315524005 | Born in South Africa |
| 315525006 | Born in South Korea |
| 315526007 | Born in Spain |
| 315527003 | Born in Sri Lanka |
| 315528008 | Born in St. Kitts and Nevis |
| 315529000 | Born in St. Lucia |
| 315530005 | Born in St. Vincent |
| 315531009 | Born in Sudan |
| 315532002 | Born in Suriname |
| 315533007 | Born in Swaziland |
| 315534001 | Born in Sweden |
| 315535000 | Born in Switzerland |
| 315536004 | Born in Syria |
| 315537008 | Born in Taiwan |
| 315538003 | Born in Tajikistan |
| 315539006 | Born in Tanzania |
| 315540008 | Born in Thailand |
| 315542000 | Born in The Netherlands |
| 315543005 | Born in Togo |
| 315544004 | Born in Tonga |
| 315545003 | Born in Trinidad and Tobago |
| 315546002 | Born in Tunisia |
| 315547006 | Born in Turkey |
| 315548001 | Born in Turkmenistan |
| 315549009 | Born in Tuvalu |
| 315550009 | Born in United States of America |
| 315551008 | Born in Uganda |
| 315552001 | Born in Ukraine |
| 315553006 | Born in United Arab Emirates |
| 315554000 | Born in Uruguay |
| 315555004 | Born in Uzbekistan |
| 315556003 | Born in Vanuatu |
| 315557007 | Born in Vatican City |
| 315558002 | Born in Venezuela |
| 315559005 | Born in Vietnam |
| 315561001 | Born in Western Samoa |
| 315562008 | Born in Yugoslavia |
| 315563003 | Born in Zaire |
| 315564009 | Born in Zambia |
| 315565005 | Born in Zimbabwe |
| 315566006 | Main spoken language Arabic |
| 315567002 | Main spoken language Bengali |
| 315568007 | Main spoken language Cantonese |
| 315569004 | Main spoken language Czech |
| 315571004 | Main spoken language French |
| 315572006 | Main spoken language Gujerati |
| 315574007 | Main spoken language Hausa |
| 315575008 | Main spoken language Hindi |
| 315576009 | Main spoken language Iba |
| 315577000 | Main spoken language Kutchi |
| 315578005 | Main spoken language Mandarin |
| 315579002 | Main spoken language Polish |
| 315580004 | Main spoken language Portuguese |
| 315581000 | Main spoken language Punjabi |
| 315582007 | Main spoken language Russian |
| 315583002 | Main spoken language Somali |
| 315584008 | Main spoken language Spanish |
| 315585009 | Main spoken language Swahili |
| 315586005 | Main spoken language Sylheti |
| 315587001 | Main spoken language Tamil |
| 315588006 | Main spoken language Urdu |
| 315589003 | Main spoken language Yoruba |
| 315590007 | Born in Yemen |
| 315593009 | Need for interpreter |
| 315594003 | Interpreter needed |
| 341651000000107 | Born in former Yugoslav Republic of Macedonia |
| 342961000000105 | Born in British overseas territory |
| 342991000000104 | Born in Montserrat |
| 343021000000109 | Born in Bermuda |
| 343711000000101 | Flemish language interpreter needed |
| 343771000000106 | Kutchi language interpreter needed |
| 345761000000103 | Born in French overseas region, department, collectivity or territory |
| 345801000000108 | Born in Martinique |
| 352901000000108 | Slovak language interpreter needed |
| 352931000000102 | Ndebele language interpreter needed |
| 353881000000101 | Romanian language interpreter needed |
| 353921000000107 | Bulgarian language interpreter needed |
| 359641000000107 | Mongolian language interpreter needed |
| 359671000000101 | Moldavian language interpreter needed |
| 359701000000102 | Marathi language interpreter needed |
| 359731000000108 | Maltese language interpreter needed |
| 359761000000103 | Luganda language interpreter needed |
| 359791000000109 | Ethiopian language interpreter needed |
| 359821000000104 | Brawa language interpreter needed |
| 359851000000109 | Kirghiz language interpreter needed |
| 359881000000103 | Iban language interpreter needed |
| 359971000000108 | Macedonian language interpreter needed |
| 360011000000101 | Malagasy language interpreter needed |
| 360071000000106 | Latvian language interpreter needed |
| 360101000000102 | Kannada language interpreter needed |
| 360131000000108 | Kinyarwanda language interpreter needed |
| 360161000000103 | Malay language interpreter needed |
| 360191000000109 | Kashmiri language interpreter needed |
| 360221000000102 | Kazakh language interpreter needed |
| 360281000000101 | Javanese language interpreter needed |
| 360311000000103 | Inuktitut language interpreter needed |
| 360341000000102 | Interlingue language interpreter needed |
| 360371000000108 | Lao language interpreter needed |
| 360431000000104 | Hungarian language interpreter needed |
| 361441000000100 | Guarani language interpreter needed |
| 361471000000106 | Georgian language interpreter needed |
| 361501000000104 | Frisian language interpreter needed |
| 361791000000100 | Faroese language interpreter needed |
| 361821000000105 | Fijian language interpreter needed |
| 361881000000106 | Esperanto language interpreter needed |
| 361911000000106 | Estonian language interpreter needed |
| 361971000000101 | Danish language interpreter needed |
| 362041000000101 | Catalan language interpreter needed |
| 362071000000107 | Belarusian language interpreter needed |
| 362111000000101 | Indonesian language interpreter needed |
| 362141000000100 | Breton language interpreter needed |
| 362171000000106 | Bislama language interpreter needed |
| 362201000000107 | Bihari language interpreter needed |
| 362231000000101 | Assamese language interpreter needed |
| 362261000000106 | Armenian language interpreter needed |
| 362371000000109 | Burmese language interpreter needed |
| 362571000000102 | Afrikaans language interpreter needed |
| 362611000000106 | Azerbaijani language interpreter needed |
| 362641000000107 | Basque language interpreter needed |
| 362691000000102 | Afar language interpreter needed |
| 362721000000106 | Abkhazian language interpreter needed |
| 362821000000104 | Zulu language interpreter needed |
| 362961000000101 | Uzbek language interpreter needed |
| 362991000000107 | Oromo language interpreter needed |
| 363021000000102 | Yiddish language interpreter needed |
| 363061000000105 | Rundi language interpreter needed |
| 363141000000105 | Tibetan language interpreter needed |
| 363181000000102 | Tsonga language interpreter needed |
| 363301000000109 | Twi language interpreter needed |
| 363341000000107 | Telugu language interpreter needed |
| 363461000000100 | Tongan language interpreter needed |
| 363551000000104 | Turkmen language interpreter needed |
| 363591000000107 | Slovenian language interpreter needed |
| 363621000000105 | Swati language interpreter needed |
| 363651000000100 | Southern Sotho language interpreter needed |
| 363681000000106 | Tajik language interpreter needed |
| 363711000000105 | Sindhi language interpreter needed |
| 363781000000103 | Sundanese language interpreter needed |
| 363841000000104 | Samoan language interpreter needed |
| 363941000000108 | Tswana language interpreter needed |
| 364011000000105 | Quechua language interpreter needed |
| 364051000000109 | Sango language interpreter needed |
| 364141000000107 | Uighur language interpreter needed |
| 364211000000103 | Oriya language interpreter needed |
| 364251000000104 | Nepali language interpreter needed |
| 364311000000108 | Occitan language interpreter needed |
| 364351000000107 | Nauru language interpreter needed |
| 364481000000106 | Romansh language interpreter needed |
| 364511000000100 | Xhosa language interpreter needed |
| 370159000 | Country of birth |
| 390790000 | Asylum seeker |
| 395108007 | Main spoken language Farsi |
| 395109004 | Main spoken language Kurdish |
| 395110009 | Main spoken language Shona |
| 407642001 | Main spoken language Italian |
| 407643006 | Main spoken language German |
| 407648002 | Main spoken language Albanian |
| 407650005 | Main spoken language Croatian |
| 407652002 | Main spoken language Greek |
| 407654001 | Main spoken language Japanese |
| 407655000 | Main spoken language Korean |
| 407656004 | Main spoken language Lithuanian |
| 407657008 | Main spoken language Turkish |
| 407659006 | Main spoken language Ukrainian |
| 407661002 | Main spoken language Vietnamese |
| 408507007 | Main spoken language Amharic |
| 408513003 | Main spoken language Brawa |
| 408514009 | Main spoken language Igbo |
| 408515005 | Main spoken language Ethiopian |
| 408516006 | Main spoken language Swedish |
| 408517002 | Main spoken language Tagalog |
| 408518007 | Main spoken language Sinhala |
| 408519004 | Main spoken language Thai |
| 408520005 | Main spoken language Flemish |
| 408521009 | Main spoken language French Créole |
| 408522002 | Main spoken language Gaelic |
| 408523007 | Main spoken language Hakka |
| 408524001 | Main spoken language Hebrew |
| 408525000 | Main spoken language Akan |
| 408526004 | Main spoken language Lingala |
| 408527008 | Main spoken language Luganda |
| 408528003 | Main spoken language Dutch |
| 408529006 | Main spoken language Malayalam |
| 408530001 | Main spoken language Norwegian |
| 408531002 | Main spoken language Pashto |
| 408533004 | Main spoken language Tigrinya |
| 408534005 | Main spoken language Patois |
| 408535006 | Main spoken language Serbian |
| 413323004 | Refugee family |
| 414640006 | Main spoken language Finnish |
| 416625007 | Family reunion immigrant |
| 423785008 | Provision of interpreter/translator services |
| 442389001 | Sign language |
| 445067002 | Country of birth unknown |
| 445075008 | Request for language interpreter service |
| 446654005 | Refugee |
| 503061000000107 | Born in Anguilla |
| 503091000000101 | Born in Dominica |
| 503511000000100 | Main spoken language Filipino |
| 511841000000102 | Main spoken language Hindko |
| 609092003 | Main spoken language Bamun |
| 609093008 | Main spoken language Dari |
| 609094002 | Main spoken language Konkani |
| 609095001 | Main spoken language Tetum |
| 698651000 | Main spoken language Abkhazian |
| 698652007 | Main spoken language Afar |
| 698653002 | Main spoken language Afrikaans |
| 698654008 | Main spoken language Basque |
| 698655009 | Main spoken language Belarusian |
| 698656005 | Main spoken language Bihari |
| 698657001 | Main spoken language Breton |
| 698658006 | Main spoken language Corsican |
| 698659003 | Main spoken language Estonian |
| 698660008 | Main spoken language Faroese |
| 698661007 | Main spoken language Frisian |
| 698662000 | Main spoken language Galician |
| 698663005 | Main spoken language Icelandic |
| 698664004 | Main spoken language Interlingua |
| 698665003 | Main spoken language Inuktitut |
| 698666002 | Main spoken language Inupiaq |
| 698667006 | Main spoken language Irish |
| 698668001 | Main spoken language Kazakh |
| 698669009 | Main spoken language Lao |
| 698670005 | Main spoken language Macedonian |
| 698671009 | Main spoken language Malagasy |
| 698672002 | Main spoken language Malay |
| 698673007 | Main spoken language Māori |
| 698674001 | Main spoken language Marathi |
| 698675000 | Main spoken language Mongolian |
| 698676004 | Main spoken language Nepali |
| 698677008 | Main spoken language Quechua |
| 698678003 | Main spoken language Romanian |
| 698679006 | Main spoken language Rundi |
| 698680009 | Main spoken language Samoan |
| 698681008 | Main spoken language Sundanese |
| 698682001 | Main spoken language Tajik |
| 698683006 | Main spoken language Tsonga |
| 698684000 | Main spoken language Tswana |
| 698685004 | Main spoken language Twi |
| 698885002 | Main spoken language Armenian |
| 698886001 | Main spoken language Assamese |
| 698887005 | Main spoken language Aymara |
| 698888000 | Main spoken language Azerbaijani |
| 698889008 | Main spoken language Bashkir |
| 698890004 | Main spoken language Bislama |
| 698891000 | Main spoken language Burmese |
| 698892007 | Main spoken language Catalan |
| 698893002 | Main spoken language Central Khmer |
| 698894008 | Main spoken language Danish |
| 698895009 | Main spoken language Jonkha |
| 698896005 | Main spoken language Esperanto |
| 698897001 | Main spoken language Fijian |
| 698898006 | Main spoken language Georgian |
| 698899003 | Main spoken language Guarani |
| 698900008 | Main spoken language Hungarian |
| 698901007 | Main spoken language Indonesian |
| 698902000 | Main spoken language Occidental |
| 698903005 | Main spoken language Javanese |
| 698904004 | Main spoken language Kalaallisut |
| 698905003 | Main spoken language Kanarese |
| 698906002 | Main spoken language Kashmiri |
| 698907006 | Main spoken language Ruanda |
| 698908001 | Main spoken language Kirgiz |
| 698909009 | Main spoken language Latvian |
| 698910004 | Main spoken language Maltese |
| 698911000 | Main spoken language Moldavian |
| 698912007 | Main spoken language Nauruan |
| 698913002 | Main spoken language Ndebele |
| 698914008 | Main spoken language Occitan |
| 698915009 | Main spoken language Oriya |
| 698916005 | Main spoken language Oromo |
| 698917001 | Main spoken language Romansh |
| 698918006 | Main spoken language Sango |
| 698919003 | Main spoken language Sindhi |
| 698920009 | Main spoken language Slovak |
| 698921008 | Main spoken language Slovenian |
| 698922001 | Main spoken language Southern Sotho |
| 698923006 | Main spoken language Swazi |
| 698924000 | Main spoken language Tatar |
| 698925004 | Main spoken language Telugu |
| 698926003 | Main spoken language Tibetan |
| 698927007 | Main spoken language Tongan |
| 698928002 | Main spoken language Turkmen |
| 698929005 | Main spoken language Uigur |
| 698930000 | Main spoken language Uzbek |
| 698932008 | Main spoken language Wolof |
| 698933003 | Main spoken language Xhosa |
| 698934009 | Main spoken language Yiddish |
| 698935005 | Main spoken language Chuang |
| 698936006 | Main spoken language Zulu |
| 699945003 | Main spoken language Bulgarian |
| 704591004 | Australian sign language |
| 704619009 | American Sign Language |
| 705023006 | Born in Democratic Republic of Congo |
| 718512007 | Main spoken language Romany |
| 728611000000100 | Asylum seeker awaiting decision on refugee status |
| 728621000000106 | Asylum seeker with application for asylum refused |
| 728631000000108 | Asylum seeker with humanitarian protection status |
| 728641000000104 | Person granted indefinite leave to remain in United Kingdom |
| 729041000000101 | Main spoken language Nyanja |
| 729051000000103 | Main spoken language Fulani |
| 729061000000100 | Main spoken language Kikuyu |
| 729851000000109 | Asylum seeker with discretionary leave to remain |
| 734998001 | Victim of human trafficking |
| 735204006 | Born in Serbia |
| 736790000 | Interpreter booked |
| 745664000 | Requires telephone language interpreter service |
| 748241000000103 | Unaccompanied child asylum seeker |
| 768761000000104 | Born in Aruba |
| 787661000000108 | Interpreter booked |
| 809341000000106 | Main spoken language Aragonese |
| 811031000000102 | Has United Kingdom student visa |
| 811111000000106 | Has United Kingdom general visitor visa |
| 841311000000103 | Country of origin high risk for blood-borne virus |
| 841321000000109 | History of medical intervention in high risk country for blood-borne virus |
| 858651000000103 | Born in Montenegro |
| 863561000000103 | Victim of human trafficking |
| 910241000000102 | First language not English |
| 918951000000105 | Born in Samoa |
| 918971000000101 | Born in Wallis and Futuna |
| 918991000000102 | Born in United States Virgin Islands |
| 919051000000101 | Born in British Virgin Islands |
| 919071000000105 | Born in Saint Vincent and the Grenadines |
| 919101000000101 | Born in United States Minor Outlying Islands |
| 919121000000105 | Born in Tokelau |
| 919161000000102 | Born in Turks and Caicos Islands |
| 919181000000106 | Born in Sint Maarten |
| 919201000000105 | Born in Saint-Martin |
| 919221000000101 | Born in Saint Pierre and Miquelon |
| 919241000000108 | Born in Saint Helena, Ascension and Tristan da Cunha |
| 919261000000109 | Born in South Georgia and the South Sandwich Islands |
| 919281000000100 | Born in French Polynesia |
| 919301000000104 | Born in French Guiana |
| 919321000000108 | Born in Falkland Islands |
| 919641000000105 | Born in Belarus |
| 919661000000106 | Born in Republic of Moldova |
| 919681000000102 | Born in American Samoa |
| 919711000000103 | Born in Bonaire, Sint Eustatius and Saba |
| 919731000000106 | Born in British Indian Ocean Territory |
| 919751000000104 | Born in Cayman Islands |
| 919771000000108 | Born in French Southern Territories |
| 919791000000107 | Born in Mayotte |
| 919811000000108 | Born in Pitcairn Islands |
| 919831000000100 | Born in Christmas Island |
| 919851000000107 | Born in Cocos (Keeling) Islands |
| 919871000000103 | Born in Cook Islands |
| 919891000000104 | Born in Guadeloupe |
| 919911000000101 | Born in Guam |
| 919931000000109 | Born in Guernsey |
| 919951000000102 | Born in Jersey |
| 919971000000106 | Born in Isle of Man |
| 919991000000105 | Born in Federated States of Micronesia |
| 920011000000104 | Born in Marshall Islands |
| 920031000000107 | Born in Niue |
| 920051000000100 | Born in Norfolk Island |
| 920071000000109 | Born in Northern Mariana Islands |
| 920091000000108 | Born in Palau |
| 920331000000109 | Born in Antarctica |
| 920521000000103 | Born in Faroe Islands |
| 920541000000105 | Born in Greenland |
| 920561000000106 | Born in Svalbard and Jan Mayen |
| 920581000000102 | Born in Åland Islands |
| 920601000000106 | Born in Réunion |
| 920761000000100 | Born in Macao |
| 920781000000109 | Born in Western Sahara |
| 920801000000105 | Born in New Caledonia |
| 923701000000106 | Born in Curaçao |
| 945731000000104 | Hands-on signing interpreter needed |
| 970441000000105 | Main spoken language Avaric |
| 970451000000108 | Main spoken language Avestan |
| 970461000000106 | Main spoken language Bambara |
| 970471000000104 | Main spoken language Bosnian |
| 970481000000102 | Main spoken language Chamorro |
| 970491000000100 | Main spoken language Chechen |
| 970501000000106 | Main spoken language Chinese |
| 970511000000108 | Main spoken language Church Slavic |
| 970521000000102 | Main spoken language Chuvash |
| 970541000000109 | Main spoken language Cree |
| 970551000000107 | Main spoken language Dhivehi |
| 970561000000105 | Main spoken language Ewe |
| 970601000000105 | Main spoken language Haitian |
| 970611000000107 | Main spoken language Herero |
| 970621000000101 | Main spoken language Hiri Motu |
| 970631000000104 | Main spoken language Ido |
| 970641000000108 | Main spoken language Kanuri |
| 970651000000106 | Main spoken language Komi |
| 970661000000109 | Main spoken language Kongo |
| 970681000000100 | Main spoken language Kuanyama |
| 970691000000103 | Main spoken language Latin |
| 970701000000103 | Main spoken language Limburgan |
| 970711000000101 | Main spoken language Luba-Katanga |
| 970721000000107 | Main spoken language Luxembourgish |
| 970751000000102 | Main spoken language Marshallese |
| 970771000000106 | Main spoken language Navajo |
| 970781000000108 | Main spoken language Ndonga |
| 970801000000109 | Main spoken language Northern Ndebele |
| 970811000000106 | Main spoken language Northern Sami |
| 970821000000100 | Main spoken language Norwegian Bokmål |
| 970831000000103 | Main spoken language Norwegian Nynorsk |
| 970851000000105 | Main spoken language Ojibwa |
| 970871000000101 | Main spoken language Ossetian |
| 970881000000104 | Main spoken language Pali |
| 970911000000104 | Main spoken language Pushto |
| 970921000000105 | Main spoken language Sanskrit |
| 970931000000107 | Main spoken language Sardinian |
| 970961000000102 | Main spoken language Nuosu |
| 970971000000109 | Main spoken language South Ndebele |
| 970991000000108 | Main spoken language Tahitian |
| 971011000000109 | Main spoken language Venda |
| 971021000000103 | Main spoken language Volapük |
| 971031000000101 | Main spoken language Walloon |
| 971041000000105 | Main spoken language Western Frisian |
| 972511000000109 | Aragonese interpreter needed |
| 972521000000103 | Avaric interpreter needed |
| 972531000000101 | Avestan interpreter needed |
| 972541000000105 | Bambara interpreter needed |
| 972551000000108 | Bashkir interpreter needed |
| 972561000000106 | Bosnian interpreter needed |
| 972571000000104 | Chamorro interpreter needed |
| 972581000000102 | Chechen interpreter needed |
| 972591000000100 | Chinese interpreter needed |
| 972601000000106 | Church Slavic interpreter needed |
| 972641000000109 | Dhivehi interpreter needed |
| 972651000000107 | Ewe interpreter needed |
| 972671000000103 | Fulah interpreter needed |
| 972691000000104 | Herero interpreter needed |
| 972711000000102 | Ido interpreter needed |
| 972721000000108 | Interlingua interpreter needed |
| 972731000000105 | Irish interpreter needed |
| 972751000000103 | Kikuyu interpreter needed |
| 972781000000109 | Kongo interpreter needed |
| 972801000000105 | Latin interpreter needed |
| 972831000000104 | Luxembourgish interpreter needed |
| 972881000000100 | Navajo interpreter needed |
| 972911000000100 | North Ndebele interpreter needed |
| 972931000000108 | Norwegian Bokmål interpreter needed |
| 972951000000101 | Nyanja interpreter needed |
| 973001000000108 | Ossetian interpreter needed |
| 973061000000107 | Serbo-Croatian interpreter needed |
| 973121000000100 | Venda interpreter needed |
| 973131000000103 | Volapük interpreter needed |
| 973141000000107 | Walloon interpreter needed |
| 973161000000108 | Wolof interpreter needed |

### Supplementary Table 2: Country-of-birth SNOMED CT codes used at least once between 2011-2024

| **SNOMED CT code** | **Description** |
| --- | --- |
| 1193634005 | Born in Cabo Verde |
| 205091000000107 | Born in Eritrea |
| 315365009 | Born in Afghanistan |
| 315366005 | Born in Albania |
| 315367001 | Born in Algeria |
| 315368006 | Born in Andorra |
| 315369003 | Born in Angola |
| 315370002 | Born in Antigua and Barbuda |
| 315372005 | Born in Argentina |
| 315373000 | Born in Armenia |
| 315374006 | Born in Australia |
| 315375007 | Born in Austria |
| 315376008 | Born in Azerbaijan |
| 315377004 | Born in Bahamas |
| 315378009 | Born in Bahrain |
| 315379001 | Born in Bangladesh |
| 315380003 | Born in Barbados |
| 315381004 | Born in Belgium |
| 315382006 | Born in Belize |
| 315383001 | Born in Belorussia |
| 315384007 | Born in Benin |
| 315385008 | Born in Bhutan |
| 315386009 | Born in Bolivia |
| 315387000 | Born in Bosnia - Herzegovnia |
| 315388005 | Born in Botswana |
| 315389002 | Born in Brazil |
| 315390006 | Born in British Guyana |
| 315391005 | Born in Brunei |
| 315392003 | Born in Bulgaria |
| 315393008 | Born in Burkina Faso |
| 315394002 | Born in Burma |
| 315395001 | Born in Burundi |
| 315396000 | Born in Cambodia |
| 315397009 | Born in Cameroon |
| 315398004 | Born in Canada |
| 315399007 | Born in Cape Verde Islands |
| 315400000 | Born in Central African Republic |
| 315401001 | Born in Chad |
| 315402008 | Born in Chechnya |
| 315403003 | Born in Chile |
| 315404009 | Born in China |
| 315405005 | Born in Colombia |
| 315406006 | Born in Comoros Islands |
| 315407002 | Born in Congo |
| 315408007 | Born in Costa Rica |
| 315409004 | Born in Croatia |
| 315410009 | Born in Cuba |
| 315411008 | Born in Cyprus |
| 315412001 | Born in Czech Republic |
| 315413006 | Born in Democratic People's Republic of Korea |
| 315414000 | Born in Denmark |
| 315415004 | Born in Djibouti |
| 315416003 | Born in Dominican Republic |
| 315417007 | Born in East Timor |
| 315418002 | Born in Ecuador |
| 315419005 | Born in Egypt |
| 315420004 | Born in El Salvador |
| 315422007 | Born in Equatorial Guinea |
| 315423002 | Born in Estonia |
| 315424008 | Born in Ethiopia |
| 315425009 | Born in Fiji |
| 315426005 | Born in Finland |
| 315427001 | Born in France |
| 315428006 | Born in Gabon |
| 315429003 | Born in Gambia |
| 315430008 | Born in Georgia |
| 315431007 | Born in Germany |
| 315432000 | Born in Ghana |
| 315433005 | Born in Greece |
| 315434004 | Born in Grenada |
| 315435003 | Born in Guatemala |
| 315436002 | Born in Guinea Bissau |
| 315437006 | Born in Guinea Republic |
| 315438001 | Born in Guyana |
| 315439009 | Born in Haiti |
| 315440006 | Born in Honduras |
| 315441005 | Born in Hong Kong |
| 315442003 | Born in Hungary |
| 315443008 | Born in Iceland |
| 315444002 | Born in India |
| 315445001 | Born in Indonesia |
| 315446000 | Born in Iran |
| 315447009 | Born in Iraq |
| 315448004 | Born in Ireland |
| 315449007 | Born in Israel |
| 315450007 | Born in Italy |
| 315451006 | Born in Ivory Coast |
| 315452004 | Born in Jamaica |
| 315453009 | Born in Japan |
| 315454003 | Born in Jordan |
| 315455002 | Born in Kazakhstan |
| 315456001 | Born in Kenya |
| 315457005 | Born in Kiribati |
| 315458000 | Born in Kosovo |
| 315459008 | Born in Kuwait |
| 315460003 | Born in Kyrgyzstan |
| 315461004 | Born in Laos |
| 315462006 | Born in Latvia |
| 315463001 | Born in Lebanon |
| 315464007 | Born in Lesotho |
| 315465008 | Born in Liberia |
| 315466009 | Born in Libya |
| 315467000 | Born in Liechtenstein |
| 315468005 | Born in Lithuania |
| 315469002 | Born in Luxembourg |
| 315470001 | Born in Madagascar |
| 315471002 | Born in Malawi |
| 315473004 | Born in Malaysia |
| 315474005 | Born in Maldives |
| 315475006 | Born in Mali |
| 315476007 | Born in Malta |
| 315477003 | Born in Mauritania |
| 315478008 | Born in Mauritius |
| 315479000 | Born in Mexico |
| 315480002 | Born in Moldavia |
| 315481003 | Born in Monaco |
| 315482005 | Born in Mongolia |
| 315483000 | Born in Morocco |
| 315484006 | Born in Mozambique |
| 315485007 | Born in Namibia |
| 315486008 | Born in Nauru |
| 315487004 | Born in Nepal |
| 315488009 | Born in New Zealand |
| 315489001 | Born in Nicaragua |
| 315490005 | Born in Niger |
| 315491009 | Born in Nigeria |
| 315492002 | Born in North Korea |
| 315494001 | Born in Norway |
| 315495000 | Born in Oman |
| 315496004 | Born in Pakistan |
| 315497008 | Born in Palestine |
| 315498003 | Born in Panama |
| 315499006 | Born in Papua New Guinea |
| 315500002 | Born in Paraguay |
| 315501003 | Born in Peru |
| 315502005 | Born in Philippines |
| 315503000 | Born in Poland |
| 315504006 | Born in Portugal |
| 315505007 | Born in Puerto Rico |
| 315506008 | Born in Qatar |
| 315507004 | Born in Republic of Ireland |
| 315508009 | Born in Republic of Korea |
| 315509001 | Born in Romania |
| 315510006 | Born in Russia |
| 315511005 | Born in Rwanda |
| 315512003 | Born in San Marino |
| 315513008 | Born in Sao Tome and Principe |
| 315514002 | Born in Saudi Arabia |
| 315516000 | Born in Senegal |
| 315517009 | Born in Seychelles |
| 315518004 | Born in Sierra Leone |
| 315519007 | Born in Singapore |
| 315520001 | Born in Slovakia |
| 315521002 | Born in Slovenia |
| 315522009 | Born in Solomon Islands |
| 315523004 | Born in Somalia |
| 315524005 | Born in South Africa |
| 315525006 | Born in South Korea |
| 315526007 | Born in Spain |
| 315527003 | Born in Sri Lanka |
| 315528008 | Born in St. Kitts and Nevis |
| 315529000 | Born in St. Lucia |
| 315530005 | Born in St. Vincent |
| 315531009 | Born in Sudan |
| 315532002 | Born in Suriname |
| 315533007 | Born in Swaziland |
| 315534001 | Born in Sweden |
| 315535000 | Born in Switzerland |
| 315536004 | Born in Syria |
| 315537008 | Born in Taiwan |
| 315538003 | Born in Tajikistan |
| 315539006 | Born in Tanzania |
| 315540008 | Born in Thailand |
| 315542000 | Born in The Netherlands |
| 315543005 | Born in Togo |
| 315544004 | Born in Tonga |
| 315545003 | Born in Trinidad and Tobago |
| 315546002 | Born in Tunisia |
| 315547006 | Born in Turkey |
| 315548001 | Born in Turkmenistan |
| 315549009 | Born in Tuvalu |
| 315550009 | Born in United States of America |
| 315551008 | Born in Uganda |
| 315552001 | Born in Ukraine |
| 315553006 | Born in United Arab Emirates |
| 315554000 | Born in Uruguay |
| 315555004 | Born in Uzbekistan |
| 315556003 | Born in Vanuatu |
| 315557007 | Born in Vatican City |
| 315558002 | Born in Venezuela |
| 315559005 | Born in Vietnam |
| 315561001 | Born in Western Samoa |
| 315562008 | Born in Yugoslavia |
| 315563003 | Born in Zaire |
| 315564009 | Born in Zambia |
| 315565005 | Born in Zimbabwe |
| 315590007 | Born in Yemen |
| 341651000000107 | Born in former Yugoslav Republic of Macedonia |
| 342961000000105 | Born in British overseas territory |
| 342991000000104 | Born in Montserrat |
| 343021000000109 | Born in Bermuda |
| 345761000000103 | Born in French overseas region, department, collectivity or territory |
| 345801000000108 | Born in Martinique |
| 503061000000107 | Born in Anguilla |
| 503091000000101 | Born in Dominica |
| 705023006 | Born in Democratic Republic of Congo |
| 735204006 | Born in Serbia |
| 768761000000104 | Born in Aruba |
| 858651000000103 | Born in Montenegro |
| 918951000000105 | Born in Samoa |
| 918971000000101 | Born in Wallis and Futuna |
| 918991000000102 | Born in United States Virgin Islands |
| 919051000000101 | Born in British Virgin Islands |
| 919071000000105 | Born in Saint Vincent and the Grenadines |
| 919101000000101 | Born in United States Minor Outlying Islands |
| 919121000000105 | Born in Tokelau |
| 919161000000102 | Born in Turks and Caicos Islands |
| 919181000000106 | Born in Sint Maarten |
| 919201000000105 | Born in Saint-Martin |
| 919221000000101 | Born in Saint Pierre and Miquelon |
| 919241000000108 | Born in Saint Helena, Ascension and Tristan da Cunha |
| 919261000000109 | Born in South Georgia and the South Sandwich Islands |
| 919281000000100 | Born in French Polynesia |
| 919301000000104 | Born in French Guiana |
| 919321000000108 | Born in Falkland Islands |
| 919641000000105 | Born in Belarus |
| 919661000000106 | Born in Republic of Moldova |
| 919681000000102 | Born in American Samoa |
| 919711000000103 | Born in Bonaire, Sint Eustatius and Saba |
| 919731000000106 | Born in British Indian Ocean Territory |
| 919751000000104 | Born in Cayman Islands |
| 919771000000108 | Born in French Southern Territories |
| 919791000000107 | Born in Mayotte |
| 919811000000108 | Born in Pitcairn Islands |
| 919831000000100 | Born in Christmas Island |
| 919851000000107 | Born in Cocos (Keeling) Islands |
| 919871000000103 | Born in Cook Islands |
| 919891000000104 | Born in Guadeloupe |
| 919911000000101 | Born in Guam |
| 919931000000109 | Born in Guernsey |
| 919951000000102 | Born in Jersey |
| 919971000000106 | Born in Isle of Man |
| 919991000000105 | Born in Federated States of Micronesia |
| 920011000000104 | Born in Marshall Islands |
| 920031000000107 | Born in Niue |
| 920051000000100 | Born in Norfolk Island |
| 920071000000109 | Born in Northern Mariana Islands |
| 920091000000108 | Born in Palau |
| 920331000000109 | Born in Antarctica |
| 920521000000103 | Born in Faroe Islands |
| 920541000000105 | Born in Greenland |
| 920561000000106 | Born in Svalbard and Jan Mayen |
| 920581000000102 | Born in Åland Islands |
| 920601000000106 | Born in Réunion |
| 920761000000100 | Born in Macao |
| 920781000000109 | Born in Western Sahara |
| 920801000000105 | Born in New Caledonia |
| 923701000000106 | Born in Curaçao |

### Supplementary Table 3: Immigration legal status SNOMED CT codes used at least once between 2011-2024

| **SNOMED CT code** | **Description** |
| --- | --- |
| 1057331000000104 | Signposting to Refugee Council |
| 1085811000000100 | History of detention in immigration removal centre |
| 1364151000000105 | Immigration Removal Centre Assessment Toolkit discharge planning screening |
| 1364221000000109 | Immigration Removal Centre Assessment Toolkit reception screening |
| 1364251000000104 | Immigration Removal Centre Assessment Toolkit reception screening declined |
| 1364261000000101 | Immigration Removal Centre Assessment Toolkit secondary screening |
| 1364271000000108 | Immigration Removal Centre Assessment Toolkit secondary screening declined |
| 1364871000000107 | Immigration Removal Centre Assessment Toolkit discharge screening |
| 1364881000000109 | Immigration Removal Centre Assessment Toolkit discharge screening declined |
| 171420007 | Examination of refugee |
| 306211000000109 | Failed asylum seeker |
| 390790000 | Asylum seeker |
| 413323004 | Refugee family |
| 416625007 | Family reunion immigrant |
| 446654005 | Refugee |
| 728611000000100 | Asylum seeker awaiting decision on refugee status |
| 728621000000106 | Asylum seeker with application for asylum refused |
| 728631000000108 | Asylum seeker with humanitarian protection status |
| 728641000000104 | Person granted indefinite leave to remain in United Kingdom |
| 729851000000109 | Asylum seeker with discretionary leave to remain |
| 748241000000103 | Unaccompanied child asylum seeker |
| 811031000000102 | Has United Kingdom student visa |
| 811111000000106 | Has United Kingdom general visitor visa |

### Supplementary Table 4: Asylum or refugee status SNOMED CT codes used at least once between 2011-2024

| **SNOMED CT code** | **Description** |
| --- | --- |
| 1057331000000104 | Signposting to Refugee Council |
| 1085811000000100 | History of detention in immigration removal centre |
| 1364151000000105 | Immigration Removal Centre Assessment Toolkit discharge planning screening |
| 1364221000000109 | Immigration Removal Centre Assessment Toolkit reception screening |
| 1364251000000104 | Immigration Removal Centre Assessment Toolkit reception screening declined |
| 1364261000000101 | Immigration Removal Centre Assessment Toolkit secondary screening |
| 1364271000000108 | Immigration Removal Centre Assessment Toolkit secondary screening declined |
| 1364871000000107 | Immigration Removal Centre Assessment Toolkit discharge screening |
| 1364881000000109 | Immigration Removal Centre Assessment Toolkit discharge screening declined |
| 171420007 | Examination of refugee |
| 306211000000109 | Failed asylum seeker |
| 390790000 | Asylum seeker |
| 413323004 | Refugee family |
| 446654005 | Refugee |
| 728611000000100 | Asylum seeker awaiting decision on refugee status |
| 728621000000106 | Asylum seeker with application for asylum refused |
| 728631000000108 | Asylum seeker with humanitarian protection status |
| 729851000000109 | Asylum seeker with discretionary leave to remain |
| 748241000000103 | Unaccompanied child asylum seeker |

### Supplementary Table 5: Language-related SNOMED CT codes used at least once between 2011-2024

| **SNOMED CT code** | **Description** |
| --- | --- |
| 1047281000000107 | Does not speak English |
| 1047301000000108 | Does not read English |
| 1047321000000104 | Romany language interpreter needed |
| 1254713008 | Requires language interpretation service to support health literacy |
| 161141008 | Bengali language |
| 161142001 | Gujarati language |
| 161143006 | Hindi language |
| 161144000 | Pashtu language |
| 161145004 | Punjabi language |
| 161146003 | Urdu language |
| 161156004 | Language difficulty |
| 203281000000104 | Akan language interpreter needed |
| 203291000000102 | Albanian language interpreter needed |
| 203301000000103 | Amharic language interpreter needed |
| 203311000000101 | Arabic language interpreter needed |
| 203321000000107 | Bengali language interpreter needed |
| 203371000000106 | Italian language interpreter needed |
| 203381000000108 | Cantonese language interpreter needed |
| 203391000000105 | Croatian language interpreter needed |
| 203401000000108 | Czech language interpreter needed |
| 203411000000105 | Dutch language interpreter needed |
| 203421000000104 | Persian language interpreter needed |
| 203441000000106 | French language interpreter needed |
| 203521000000103 | French Creole language interpreter needed |
| 203531000000101 | Igbo language interpreter needed |
| 203581000000102 | German language interpreter needed |
| 203591000000100 | Greek language interpreter needed |
| 203601000000106 | Gujarati language interpreter needed |
| 203611000000108 | Hakka language interpreter needed |
| 203631000000100 | Hausa language interpreter needed |
| 203641000000109 | Hebrew language interpreter needed |
| 203651000000107 | Hindi language interpreter needed |
| 203681000000101 | Japanese language interpreter needed |
| 203691000000104 | Korean language interpreter needed |
| 203701000000104 | Kurdish language interpreter needed |
| 203711000000102 | Lingala language interpreter needed |
| 203721000000108 | Lithuanian language interpreter needed |
| 203801000000105 | Ganda language interpreter needed |
| 203811000000107 | Malayalam language interpreter needed |
| 203821000000101 | Mandarin language interpreter needed |
| 203831000000104 | Norwegian language interpreter needed |
| 203841000000108 | Pashto language interpreter needed |
| 203901000000102 | Polish language interpreter needed |
| 203911000000100 | Portuguese language interpreter needed |
| 203961000000103 | Panjabi language interpreter needed |
| 203971000000105 | Russian language interpreter needed |
| 203981000000107 | Serbian language interpreter needed |
| 203991000000109 | Shona language interpreter needed |
| 204011000000102 | Sinhala language interpreter needed |
| 204021000000108 | Somali language interpreter needed |
| 204031000000105 | Spanish language interpreter needed |
| 204041000000101 | Swahili language interpreter needed |
| 204051000000103 | Swedish language interpreter needed |
| 204081000000109 | Sylheti language interpreter needed |
| 204111000000101 | Tagalog language interpreter needed |
| 204131000000109 | Tamil language interpreter needed |
| 204151000000102 | Thai language interpreter needed |
| 204171000000106 | Tigrinya language interpreter needed |
| 204191000000105 | Turkish language interpreter needed |
| 204201000000107 | Ukrainian language interpreter needed |
| 204211000000109 | Urdu language interpreter needed |
| 204221000000103 | Vietnamese language interpreter needed |
| 204241000000105 | Yoruba language interpreter needed |
| 224628009 | Hinko language |
| 297289008 | World languages |
| 297290004 | Afro-Asiatic language |
| 297291000 | Berber language |
| 297292007 | Kabyle language |
| 297296005 | Tamazight language |
| 297298006 | Chadic language |
| 297299003 | Hausa language |
| 297300006 | Cushitic language |
| 297301005 | Afar language |
| 297303008 | Oromo language |
| 297304002 | Saho language |
| 297305001 | Sidamo language |
| 297306000 | Somali language |
| 297308004 | Egyptian language |
| 297309007 | Coptic language |
| 297310002 | Semitic language |
| 297311003 | Aramaic language |
| 297312005 | Assyrian language |
| 297313000 | Syriac language |
| 297314006 | Canaanitic language |
| 297315007 | Hebrew language |
| 297316008 | Ethiopic language |
| 297317004 | Amharic language |
| 297319001 | Gurage language |
| 297320007 | Harari language |
| 297321006 | Tigre language |
| 297322004 | Tigrinya language |
| 297323009 | North Arabic language |
| 297324003 | Arabic language |
| 297325002 | Classical Arabic language |
| 297326001 | South Arabic language |
| 297327005 | Maltese language |
| 297328000 | Altaic language |
| 297329008 | Mongolian language |
| 297332006 | Tungusic language |
| 297337000 | Manchu language |
| 297339002 | Sibo language |
| 297340000 | Turkic language |
| 297341001 | Chuvash language |
| 297344009 | Khakass language |
| 297345005 | Tuvinian language |
| 297347002 | Balkar language |
| 297349004 | Kara-Kalpak language |
| 297350004 | Karachai language |
| 297351000 | Kazakh language |
| 297352007 | Kirgiz language |
| 297353002 | Kumyk language |
| 297355009 | Tatar language |
| 297358006 | Salar language |
| 297359003 | Uigur language |
| 297360008 | Uzbek language |
| 297362000 | Azerbaijani language |
| 297363005 | Turkish language |
| 297364004 | Turkmen language |
| 297367006 | Esperanto language |
| 297368001 | Interlingua language |
| 297370005 | Australian language |
| 297371009 | Aranda language |
| 297373007 | Caucasian language |
| 297375000 | Avar language |
| 297376004 | Dargin language |
| 297378003 | Lezgin language |
| 297380009 | Eastern Caucasian language |
| 297381008 | Chechen language |
| 297382001 | Ingush language |
| 297383006 | Southern Caucasian language |
| 297384000 | Georgian language |
| 297385004 | Western Caucasian language |
| 297389005 | Circassian language |
| 297391002 | Central and South American Indian language |
| 297392009 | Andean equatorial language |
| 297395006 | Aymara language |
| 297396007 | Guarani language |
| 297398008 | Quechua language |
| 297400007 | Ge-Pano-Carib language |
| 297401006 | Carib language |
| 297405002 | Tacana language |
| 297407005 | Bribri language |
| 297408000 | Cabecar language |
| 297411004 | Lenca language |
| 297414007 | Central Sudanic language |
| 297415008 | Lugbara language |
| 297416009 | Madi language |
| 297417000 | Mangbetu language |
| 297418005 | Sara language |
| 297419002 | Eastern Sudanic language |
| 297420008 | Nilotic language |
| 297421007 | Eastern Nilotic language |
| 297422000 | Bari language |
| 297424004 | Lotuko language |
| 297425003 | Masai language |
| 297426002 | Teso language |
| 297429009 | Kalenjin language |
| 297430004 | Suk language |
| 297431000 | Western Nilotic language |
| 297432007 | Acholi language |
| 297434008 | Dinka language |
| 297435009 | Lango language |
| 297436005 | Luo language |
| 297437001 | Nuer language |
| 297439003 | Nubian language |
| 297440001 | Djerma language |
| 297441002 | Dravidian language |
| 297442009 | Brahui language |
| 297444005 | Kanarese language |
| 297446007 | Kurukh language |
| 297447003 | Malayalam language |
| 297448008 | Tamil language |
| 297449000 | Telugu language |
| 297450000 | Tulu language |
| 297454009 | Fur language |
| 297455005 | Independent language |
| 297457002 | Barushaski language |
| 297459004 | Basque language |
| 297460009 | Japanese language |
| 297461008 | Korean language |
| 297463006 | Vietnamese language |
| 297464000 | Indo-European language |
| 297465004 | Albanian language |
| 297466003 | Armenian language |
| 297467007 | Baltic language |
| 297468002 | Latvian language |
| 297469005 | Lithuanian language |
| 297475001 | Irish Gaelic language |
| 297477009 | Germanic language |
| 297478004 | Northern Germanic language |
| 297479007 | Danish language |
| 297480005 | Faroese language |
| 297481009 | Icelandic language |
| 297482002 | Norwegian language |
| 297483007 | Swedish language |
| 297484001 | Western Germanic language |
| 297485000 | Afrikaans language |
| 297486004 | Dutch language |
| 297488003 | Flemish language |
| 297490002 | German language |
| 297491003 | Luxembourgian language |
| 297492005 | Yiddish language |
| 297493000 | Hellenic language |
| 297494006 | Greek language |
| 297495007 | Italic language |
| 297496008 | Latin language |
| 297497004 | Romance language |
| 297498009 | Catalan language |
| 297499001 | French language |
| 297501009 | Swiss French dialect |
| 297502002 | Italian language |
| 297503007 | Moldavian language |
| 297504001 | Portuguese language |
| 297505000 | Galician Portuguese dialect |
| 297507008 | Rhaeto-Romanic language |
| 297508003 | Rumanian language |
| 297509006 | Sardinian language |
| 297510001 | Spanish language |
| 297511002 | Slavic language |
| 297512009 | Eastern Slavic language |
| 297513004 | Belorussian language |
| 297514005 | Russian language |
| 297515006 | Ukrainian language |
| 297516007 | Southern Slavic language |
| 297517003 | Bulgarian language |
| 297518008 | Macedonian language |
| 297519000 | Serbo-Croatian language |
| 297520006 | Slovenian language |
| 297521005 | Western Slavic language |
| 297522003 | Czech language |
| 297523008 | Polish language |
| 297524002 | Slovak language |
| 297525001 | Sorbian language |
| 297526000 | Indo-Iranian language |
| 297527009 | Indic language |
| 297528004 | Assamese language |
| 297530002 | Sylhety |
| 297531003 | Bihari language |
| 297532005 | Hindustani language |
| 297533000 | Kashmiri language |
| 297534006 | Konkani language |
| 297535007 | Maldivian language |
| 297536008 | Marathi language |
| 297537004 | Nepali language |
| 297538009 | Oriya language |
| 297539001 | Pakistani punjabi language |
| 297540004 | Lehnda punjabi language |
| 297541000 | Sikh punjabi language |
| 297542007 | Rajasthani language |
| 297543002 | Romany language |
| 297544008 | Sanskrit language |
| 297545009 | Sindhi language |
| 297546005 | Sinhalese language |
| 297547001 | Iranian language |
| 297548006 | Baluchi language |
| 297549003 | Gilaki language |
| 297550003 | Kurdish language |
| 297551004 | Mazanderani language |
| 297552006 | Ossetian language |
| 297553001 | Persian language |
| 297554007 | Tajik language |
| 297555008 | Khoisan language |
| 297556009 | Bushman language |
| 297557000 | Hatsa language |
| 297561006 | Maban language |
| 297562004 | Maba language |
| 297563009 | Malayo-Polynesian language |
| 297564003 | Indonesian language |
| 297565002 | Achinese language |
| 297566001 | Balinese language |
| 297568000 | Bikol language |
| 297569008 | Buginese language |
| 297570009 | Cham language |
| 297571008 | Chamorro language |
| 297573006 | Igorot language |
| 297575004 | Javanese language |
| 297576003 | Madurese language |
| 297577007 | Malagasy language |
| 297578002 | Malay language |
| 297580008 | Minangkabau language |
| 297582000 | Pampangan language |
| 297583005 | Pangasinan language |
| 297584004 | Rhade language |
| 297585003 | Sundanese language |
| 297586002 | Tagalog language |
| 297587006 | Visayan language |
| 297592008 | Ponapean language |
| 297593003 | Trukese language |
| 297595005 | Melanesian language |
| 297596006 | Fijian language |
| 297598007 | Tolai language |
| 297600001 | Polynesian language |
| 297601002 | Hawaiian language |
| 297602009 | Māori language |
| 297603004 | Marquesan language |
| 297604005 | Niuean language |
| 297606007 | Samoan language |
| 297607003 | Tahitian language |
| 297608008 | Tongan language |
| 297612002 | Bahnar language |
| 297613007 | Khasi language |
| 297614001 | Khmer language |
| 297615000 | Mon language |
| 297620000 | Munda language |
| 297624009 | Santali language |
| 297625005 | Savara language |
| 297626006 | Niger-Congo language |
| 297627002 | Adamawa-Eastern language |
| 297628007 | Adamawa language |
| 297630009 | Eastern language (Niger-Congo) |
| 297631008 | Banda language |
| 297632001 | Gbaya language |
| 297633006 | Sango language |
| 297634000 | Zande language |
| 297635004 | Benue-Congo language |
| 297637007 | Bemba language |
| 297639005 | Bulu language |
| 297640007 | Chagga language |
| 297641006 | Chiga language |
| 297643009 | Duala language |
| 297645002 | Ganda language |
| 297646001 | Gisu language |
| 297648000 | Herero language |
| 297649008 | Kamba language |
| 297650008 | Kikuyu language |
| 297651007 | Kisii language |
| 297652000 | Kongo language |
| 297653005 | Lingala language |
| 297655003 | Lozi language |
| 297657006 | Luhya language |
| 297658001 | Lunda language |
| 297664008 | Meru language |
| 297666005 | Ndebele language |
| 297667001 | Ngala language |
| 297670002 | Nyanja language |
| 297672005 | Pedi language |
| 297673000 | Ruanda language |
| 297674006 | Rundi language |
| 297675007 | Shona language |
| 297676008 | Sotho language |
| 297678009 | Swahili language |
| 297679001 | Swazi language |
| 297681004 | Toro language |
| 297682006 | Tsonga language |
| 297683001 | Tswana language |
| 297684007 | Tumbuka language |
| 297685008 | Venda language |
| 297686009 | Xhosa language |
| 297687000 | Yao language - Bantu |
| 297689002 | Zulu language |
| 297690006 | Efik language |
| 297691005 | Ibibio language |
| 297692003 | Tiv language |
| 297693008 | Ijo language |
| 297694002 | Western Sudanic language |
| 297695001 | Gur language |
| 297696000 | Bariba language |
| 297697009 | Dagomba language |
| 297698004 | Gurma language |
| 297702000 | Kwa language |
| 297703005 | Adangme language |
| 297705003 | Bassa language |
| 297707006 | Edo language |
| 297708001 | Ewe language |
| 297710004 | Fanti language |
| 297711000 | Fon language |
| 297712007 | Ga language |
| 297714008 | Grebo language |
| 297715009 | Ibo language |
| 297716005 | Idoma language |
| 297717001 | Kru language |
| 297718006 | Nupe language |
| 297719003 | Twi language |
| 297720009 | Urhobo language |
| 297721008 | Yoruba language |
| 297722001 | Mande language |
| 297723006 | Bambara language |
| 297726003 | Loma language |
| 297727007 | Malinke language |
| 297728002 | Mende language |
| 297729005 | Soninke language |
| 297730000 | Susu language |
| 297733003 | Balante language |
| 297735005 | Fulani language |
| 297736006 | Gola language |
| 297737002 | Kissi language |
| 297738007 | Serer language |
| 297739004 | Temne language |
| 297740002 | Wolof language |
| 297741003 | Native North American language |
| 297742005 | Algonkian language |
| 297745007 | Cheyenne language |
| 297746008 | Cree language |
| 297747004 | Delaware language |
| 297748009 | Fox language |
| 297752009 | Ojibwa language |
| 297753004 | Passamaquoddy language |
| 297757003 | Chilcotin language |
| 297758008 | Chipewyan language |
| 297769006 | Seneca language |
| 297771006 | Mayan language |
| 297777005 | Mam language |
| 297778000 | Maya language |
| 297782003 | Muskogean language |
| 297783008 | Chickasaw language |
| 297785001 | Creek language |
| 297787009 | Oto-Manguean language |
| 297788004 | Chinantec language |
| 297792006 | Otomi language |
| 297794007 | Penutian language |
| 297796009 | Nez Perce language |
| 297800003 | Flathead language |
| 297801004 | Lillooet language |
| 297808005 | Omaha language |
| 297809002 | Osage language |
| 297812004 | Tanoan language |
| 297813009 | Tewa language |
| 297814003 | Tiwa language |
| 297823000 | Mayo language |
| 297824006 | Nahuatl language |
| 297832003 | Kwakiutl language |
| 297838004 | Mixe language |
| 297841008 | Zunian language |
| 297844000 | Chukchi language |
| 297849005 | Yukagir language |
| 297850005 | Papuan language |
| 297853007 | Enga language |
| 297854001 | Hagen language |
| 297855000 | Kate language |
| 297856004 | Marind language |
| 297858003 | Pidgin and Créole language |
| 297860001 | French Créole language |
| 297862009 | Krio language |
| 297863004 | Papiamento language |
| 297864005 | Pidgin English language |
| 297865006 | Police Motu language |
| 297867003 | Saramacca language |
| 297869000 | Saharan language |
| 297870004 | Kanuri language |
| 297872007 | Sino-Tibetan language |
| 297873002 | Miao-Yao language |
| 297875009 | Sinitic language |
| 297876005 | Chinese language |
| 297877001 | Cantonese Chinese dialect |
| 297878006 | Fukienese dialect |
| 297881001 | Hakka dialect |
| 297883003 | Mandarin dialect |
| 297885005 | Tai language |
| 297887002 | Lao language |
| 297889004 | Puyi language |
| 297890008 | Shan language |
| 297891007 | Thai language |
| 297893005 | Tibeto-Burman language |
| 297894004 | Bodo language |
| 297895003 | Burmese language |
| 297896002 | Chin language |
| 297897006 | Garo language |
| 297899009 | Kachin language |
| 297900004 | Karen language |
| 297901000 | Lahu language |
| 297902007 | Lepcha language |
| 297904008 | Lushei language |
| 297905009 | Meithei language |
| 297907001 | Nakhi language |
| 297908006 | Newari language |
| 297909003 | Tibetan language |
| 297910008 | Yi language |
| 297911007 | Songhai language |
| 297912000 | Uralic language |
| 297914004 | Finnic language |
| 297915003 | Estonian language |
| 297916002 | Finnish language |
| 297920003 | Mordvin language |
| 297923001 | Hungarian language |
| 297927000 | Enets language |
| 297928005 | Nenets language |
| 297931006 | Language commonly spoken in Europe |
| 297934003 | Swiss German dialect |
| 298018001 | Kashubian Polish dialect |
| 303601000000100 | Telephone interpreting service used |
| 313421002 | Créole language |
| 313422009 | Mirpuri language |
| 314430004 | Presence of interpreter |
| 314431000 | Interpreter present |
| 315566006 | Main spoken language Arabic |
| 315567002 | Main spoken language Bengali |
| 315568007 | Main spoken language Cantonese |
| 315569004 | Main spoken language Czech |
| 315571004 | Main spoken language French |
| 315572006 | Main spoken language Gujerati |
| 315574007 | Main spoken language Hausa |
| 315575008 | Main spoken language Hindi |
| 315576009 | Main spoken language Iba |
| 315577000 | Main spoken language Kutchi |
| 315578005 | Main spoken language Mandarin |
| 315579002 | Main spoken language Polish |
| 315580004 | Main spoken language Portuguese |
| 315581000 | Main spoken language Punjabi |
| 315582007 | Main spoken language Russian |
| 315583002 | Main spoken language Somali |
| 315584008 | Main spoken language Spanish |
| 315585009 | Main spoken language Swahili |
| 315586005 | Main spoken language Sylheti |
| 315587001 | Main spoken language Tamil |
| 315588006 | Main spoken language Urdu |
| 315589003 | Main spoken language Yoruba |
| 315593009 | Need for interpreter |
| 315594003 | Interpreter needed |
| 343711000000101 | Flemish language interpreter needed |
| 343771000000106 | Kutchi language interpreter needed |
| 352901000000108 | Slovak language interpreter needed |
| 352931000000102 | Ndebele language interpreter needed |
| 353881000000101 | Romanian language interpreter needed |
| 353921000000107 | Bulgarian language interpreter needed |
| 359641000000107 | Mongolian language interpreter needed |
| 359671000000101 | Moldavian language interpreter needed |
| 359701000000102 | Marathi language interpreter needed |
| 359731000000108 | Maltese language interpreter needed |
| 359761000000103 | Luganda language interpreter needed |
| 359791000000109 | Ethiopian language interpreter needed |
| 359821000000104 | Brawa language interpreter needed |
| 359851000000109 | Kirghiz language interpreter needed |
| 359881000000103 | Iban language interpreter needed |
| 359971000000108 | Macedonian language interpreter needed |
| 360011000000101 | Malagasy language interpreter needed |
| 360071000000106 | Latvian language interpreter needed |
| 360101000000102 | Kannada language interpreter needed |
| 360131000000108 | Kinyarwanda language interpreter needed |
| 360161000000103 | Malay language interpreter needed |
| 360191000000109 | Kashmiri language interpreter needed |
| 360221000000102 | Kazakh language interpreter needed |
| 360281000000101 | Javanese language interpreter needed |
| 360311000000103 | Inuktitut language interpreter needed |
| 360341000000102 | Interlingue language interpreter needed |
| 360371000000108 | Lao language interpreter needed |
| 360431000000104 | Hungarian language interpreter needed |
| 361441000000100 | Guarani language interpreter needed |
| 361471000000106 | Georgian language interpreter needed |
| 361501000000104 | Frisian language interpreter needed |
| 361791000000100 | Faroese language interpreter needed |
| 361821000000105 | Fijian language interpreter needed |
| 361881000000106 | Esperanto language interpreter needed |
| 361911000000106 | Estonian language interpreter needed |
| 361971000000101 | Danish language interpreter needed |
| 362041000000101 | Catalan language interpreter needed |
| 362071000000107 | Belarusian language interpreter needed |
| 362111000000101 | Indonesian language interpreter needed |
| 362141000000100 | Breton language interpreter needed |
| 362171000000106 | Bislama language interpreter needed |
| 362201000000107 | Bihari language interpreter needed |
| 362231000000101 | Assamese language interpreter needed |
| 362261000000106 | Armenian language interpreter needed |
| 362371000000109 | Burmese language interpreter needed |
| 362571000000102 | Afrikaans language interpreter needed |
| 362611000000106 | Azerbaijani language interpreter needed |
| 362641000000107 | Basque language interpreter needed |
| 362691000000102 | Afar language interpreter needed |
| 362721000000106 | Abkhazian language interpreter needed |
| 362821000000104 | Zulu language interpreter needed |
| 362961000000101 | Uzbek language interpreter needed |
| 362991000000107 | Oromo language interpreter needed |
| 363021000000102 | Yiddish language interpreter needed |
| 363061000000105 | Rundi language interpreter needed |
| 363141000000105 | Tibetan language interpreter needed |
| 363181000000102 | Tsonga language interpreter needed |
| 363301000000109 | Twi language interpreter needed |
| 363341000000107 | Telugu language interpreter needed |
| 363461000000100 | Tongan language interpreter needed |
| 363551000000104 | Turkmen language interpreter needed |
| 363591000000107 | Slovenian language interpreter needed |
| 363621000000105 | Swati language interpreter needed |
| 363651000000100 | Southern Sotho language interpreter needed |
| 363681000000106 | Tajik language interpreter needed |
| 363711000000105 | Sindhi language interpreter needed |
| 363781000000103 | Sundanese language interpreter needed |
| 363841000000104 | Samoan language interpreter needed |
| 363941000000108 | Tswana language interpreter needed |
| 364011000000105 | Quechua language interpreter needed |
| 364051000000109 | Sango language interpreter needed |
| 364141000000107 | Uighur language interpreter needed |
| 364211000000103 | Oriya language interpreter needed |
| 364251000000104 | Nepali language interpreter needed |
| 364311000000108 | Occitan language interpreter needed |
| 364351000000107 | Nauru language interpreter needed |
| 364481000000106 | Romansh language interpreter needed |
| 364511000000100 | Xhosa language interpreter needed |
| 370157003 | Main spoken language |
| 395108007 | Main spoken language Farsi |
| 395109004 | Main spoken language Kurdish |
| 395110009 | Main spoken language Shona |
| 407642001 | Main spoken language Italian |
| 407643006 | Main spoken language German |
| 407648002 | Main spoken language Albanian |
| 407650005 | Main spoken language Croatian |
| 407652002 | Main spoken language Greek |
| 407654001 | Main spoken language Japanese |
| 407655000 | Main spoken language Korean |
| 407656004 | Main spoken language Lithuanian |
| 407657008 | Main spoken language Turkish |
| 407659006 | Main spoken language Ukrainian |
| 407661002 | Main spoken language Vietnamese |
| 408507007 | Main spoken language Amharic |
| 408513003 | Main spoken language Brawa |
| 408514009 | Main spoken language Igbo |
| 408515005 | Main spoken language Ethiopian |
| 408516006 | Main spoken language Swedish |
| 408517002 | Main spoken language Tagalog |
| 408518007 | Main spoken language Sinhala |
| 408519004 | Main spoken language Thai |
| 408520005 | Main spoken language Flemish |
| 408521009 | Main spoken language French Créole |
| 408523007 | Main spoken language Hakka |
| 408524001 | Main spoken language Hebrew |
| 408525000 | Main spoken language Akan |
| 408526004 | Main spoken language Lingala |
| 408527008 | Main spoken language Luganda |
| 408528003 | Main spoken language Dutch |
| 408529006 | Main spoken language Malayalam |
| 408530001 | Main spoken language Norwegian |
| 408531002 | Main spoken language Pashto |
| 408533004 | Main spoken language Tigrinya |
| 408534005 | Main spoken language Patois |
| 408535006 | Main spoken language Serbian |
| 414640006 | Main spoken language Finnish |
| 426201006 | Interpreter not available |
| 442389001 | Sign language |
| 445075008 | Request for language interpreter service |
| 503511000000100 | Main spoken language Filipino |
| 511841000000102 | Main spoken language Hindko |
| 609092003 | Main spoken language Bamun |
| 609093008 | Main spoken language Dari |
| 609094002 | Main spoken language Konkani |
| 609095001 | Main spoken language Tetum |
| 698651000 | Main spoken language Abkhazian |
| 698652007 | Main spoken language Afar |
| 698653002 | Main spoken language Afrikaans |
| 698654008 | Main spoken language Basque |
| 698655009 | Main spoken language Belarusian |
| 698656005 | Main spoken language Bihari |
| 698657001 | Main spoken language Breton |
| 698658006 | Main spoken language Corsican |
| 698659003 | Main spoken language Estonian |
| 698660008 | Main spoken language Faroese |
| 698661007 | Main spoken language Frisian |
| 698662000 | Main spoken language Galician |
| 698663005 | Main spoken language Icelandic |
| 698664004 | Main spoken language Interlingua |
| 698665003 | Main spoken language Inuktitut |
| 698666002 | Main spoken language Inupiaq |
| 698667006 | Main spoken language Irish |
| 698668001 | Main spoken language Kazakh |
| 698669009 | Main spoken language Lao |
| 698670005 | Main spoken language Macedonian |
| 698671009 | Main spoken language Malagasy |
| 698672002 | Main spoken language Malay |
| 698673007 | Main spoken language Māori |
| 698674001 | Main spoken language Marathi |
| 698675000 | Main spoken language Mongolian |
| 698676004 | Main spoken language Nepali |
| 698677008 | Main spoken language Quechua |
| 698678003 | Main spoken language Romanian |
| 698679006 | Main spoken language Rundi |
| 698680009 | Main spoken language Samoan |
| 698681008 | Main spoken language Sundanese |
| 698682001 | Main spoken language Tajik |
| 698683006 | Main spoken language Tsonga |
| 698684000 | Main spoken language Tswana |
| 698685004 | Main spoken language Twi |
| 698885002 | Main spoken language Armenian |
| 698886001 | Main spoken language Assamese |
| 698887005 | Main spoken language Aymara |
| 698888000 | Main spoken language Azerbaijani |
| 698889008 | Main spoken language Bashkir |
| 698890004 | Main spoken language Bislama |
| 698891000 | Main spoken language Burmese |
| 698892007 | Main spoken language Catalan |
| 698893002 | Main spoken language Central Khmer |
| 698894008 | Main spoken language Danish |
| 698895009 | Main spoken language Jonkha |
| 698896005 | Main spoken language Esperanto |
| 698897001 | Main spoken language Fijian |
| 698898006 | Main spoken language Georgian |
| 698899003 | Main spoken language Guarani |
| 698900008 | Main spoken language Hungarian |
| 698901007 | Main spoken language Indonesian |
| 698902000 | Main spoken language Occidental |
| 698903005 | Main spoken language Javanese |
| 698904004 | Main spoken language Kalaallisut |
| 698905003 | Main spoken language Kanarese |
| 698906002 | Main spoken language Kashmiri |
| 698907006 | Main spoken language Ruanda |
| 698908001 | Main spoken language Kirgiz |
| 698909009 | Main spoken language Latvian |
| 698910004 | Main spoken language Maltese |
| 698911000 | Main spoken language Moldavian |
| 698912007 | Main spoken language Nauruan |
| 698913002 | Main spoken language Ndebele |
| 698914008 | Main spoken language Occitan |
| 698915009 | Main spoken language Oriya |
| 698916005 | Main spoken language Oromo |
| 698917001 | Main spoken language Romansh |
| 698918006 | Main spoken language Sango |
| 698919003 | Main spoken language Sindhi |
| 698920009 | Main spoken language Slovak |
| 698921008 | Main spoken language Slovenian |
| 698922001 | Main spoken language Southern Sotho |
| 698923006 | Main spoken language Swazi |
| 698924000 | Main spoken language Tatar |
| 698925004 | Main spoken language Telugu |
| 698926003 | Main spoken language Tibetan |
| 698927007 | Main spoken language Tongan |
| 698928002 | Main spoken language Turkmen |
| 698929005 | Main spoken language Uigur |
| 698930000 | Main spoken language Uzbek |
| 698932008 | Main spoken language Wolof |
| 698933003 | Main spoken language Xhosa |
| 698934009 | Main spoken language Yiddish |
| 698935005 | Main spoken language Chuang |
| 698936006 | Main spoken language Zulu |
| 699945003 | Main spoken language Bulgarian |
| 704591004 | Australian sign language |
| 704619009 | American Sign Language |
| 718512007 | Main spoken language Romany |
| 729041000000101 | Main spoken language Nyanja |
| 729051000000103 | Main spoken language Fulani |
| 729061000000100 | Main spoken language Kikuyu |
| 736790000 | Interpreter booked |
| 787661000000108 | Interpreter booked |
| 809341000000106 | Main spoken language Aragonese |
| 970441000000105 | Main spoken language Avaric |
| 970451000000108 | Main spoken language Avestan |
| 970461000000106 | Main spoken language Bambara |
| 970471000000104 | Main spoken language Bosnian |
| 970481000000102 | Main spoken language Chamorro |
| 970491000000100 | Main spoken language Chechen |
| 970501000000106 | Main spoken language Chinese |
| 970511000000108 | Main spoken language Church Slavic |
| 970521000000102 | Main spoken language Chuvash |
| 970541000000109 | Main spoken language Cree |
| 970551000000107 | Main spoken language Dhivehi |
| 970561000000105 | Main spoken language Ewe |
| 970601000000105 | Main spoken language Haitian |
| 970611000000107 | Main spoken language Herero |
| 970621000000101 | Main spoken language Hiri Motu |
| 970631000000104 | Main spoken language Ido |
| 970641000000108 | Main spoken language Kanuri |
| 970651000000106 | Main spoken language Komi |
| 970661000000109 | Main spoken language Kongo |
| 970681000000100 | Main spoken language Kuanyama |
| 970691000000103 | Main spoken language Latin |
| 970701000000103 | Main spoken language Limburgan |
| 970711000000101 | Main spoken language Luba-Katanga |
| 970721000000107 | Main spoken language Luxembourgish |
| 970751000000102 | Main spoken language Marshallese |
| 970771000000106 | Main spoken language Navajo |
| 970781000000108 | Main spoken language Ndonga |
| 970801000000109 | Main spoken language Northern Ndebele |
| 970811000000106 | Main spoken language Northern Sami |
| 970821000000100 | Main spoken language Norwegian Bokmål |
| 970831000000103 | Main spoken language Norwegian Nynorsk |
| 970851000000105 | Main spoken language Ojibwa |
| 970871000000101 | Main spoken language Ossetian |
| 970881000000104 | Main spoken language Pali |
| 970911000000104 | Main spoken language Pushto |
| 970921000000105 | Main spoken language Sanskrit |
| 970931000000107 | Main spoken language Sardinian |
| 970961000000102 | Main spoken language Nuosu |
| 970971000000109 | Main spoken language South Ndebele |
| 970991000000108 | Main spoken language Tahitian |
| 971011000000109 | Main spoken language Venda |
| 971021000000103 | Main spoken language Volapük |
| 971031000000101 | Main spoken language Walloon |
| 971041000000105 | Main spoken language Western Frisian |
| 972511000000109 | Aragonese interpreter needed |
| 972521000000103 | Avaric interpreter needed |
| 972531000000101 | Avestan interpreter needed |
| 972541000000105 | Bambara interpreter needed |
| 972551000000108 | Bashkir interpreter needed |
| 972561000000106 | Bosnian interpreter needed |
| 972571000000104 | Chamorro interpreter needed |
| 972581000000102 | Chechen interpreter needed |
| 972591000000100 | Chinese interpreter needed |
| 972601000000106 | Church Slavic interpreter needed |
| 972641000000109 | Dhivehi interpreter needed |
| 972651000000107 | Ewe interpreter needed |
| 972671000000103 | Fulah interpreter needed |
| 972691000000104 | Herero interpreter needed |
| 972711000000102 | Ido interpreter needed |
| 972721000000108 | Interlingua interpreter needed |
| 972731000000105 | Irish interpreter needed |
| 972751000000103 | Kikuyu interpreter needed |
| 972781000000109 | Kongo interpreter needed |
| 972801000000105 | Latin interpreter needed |
| 972831000000104 | Luxembourgish interpreter needed |
| 972881000000100 | Navajo interpreter needed |
| 972911000000100 | North Ndebele interpreter needed |
| 972931000000108 | Norwegian Bokmål interpreter needed |
| 972951000000101 | Nyanja interpreter needed |
| 973001000000108 | Ossetian interpreter needed |
| 973061000000107 | Serbo-Croatian interpreter needed |
| 973121000000100 | Venda interpreter needed |
| 973131000000103 | Volapük interpreter needed |
| 973141000000107 | Walloon interpreter needed |
| 973161000000108 | Wolof interpreter needed |

### Supplementary Table 6: Interpreter need SNOMED CT codes used at least once between 2011-2024

| **SNOMED CT code** | **Description** |
| --- | --- |
| 1047281000000107 | Does not speak English |
| 1047301000000108 | Does not read English |
| 1047321000000104 | Romany language interpreter needed |
| 1254713008 | Requires language interpretation service to support health literacy |
| 203281000000104 | Akan language interpreter needed |
| 203291000000102 | Albanian language interpreter needed |
| 203301000000103 | Amharic language interpreter needed |
| 203311000000101 | Arabic language interpreter needed |
| 203321000000107 | Bengali language interpreter needed |
| 203371000000106 | Italian language interpreter needed |
| 203381000000108 | Cantonese language interpreter needed |
| 203391000000105 | Croatian language interpreter needed |
| 203401000000108 | Czech language interpreter needed |
| 203411000000105 | Dutch language interpreter needed |
| 203421000000104 | Persian language interpreter needed |
| 203441000000106 | French language interpreter needed |
| 203521000000103 | French Creole language interpreter needed |
| 203531000000101 | Igbo language interpreter needed |
| 203581000000102 | German language interpreter needed |
| 203591000000100 | Greek language interpreter needed |
| 203601000000106 | Gujarati language interpreter needed |
| 203611000000108 | Hakka language interpreter needed |
| 203631000000100 | Hausa language interpreter needed |
| 203641000000109 | Hebrew language interpreter needed |
| 203651000000107 | Hindi language interpreter needed |
| 203681000000101 | Japanese language interpreter needed |
| 203691000000104 | Korean language interpreter needed |
| 203701000000104 | Kurdish language interpreter needed |
| 203711000000102 | Lingala language interpreter needed |
| 203721000000108 | Lithuanian language interpreter needed |
| 203801000000105 | Ganda language interpreter needed |
| 203811000000107 | Malayalam language interpreter needed |
| 203821000000101 | Mandarin language interpreter needed |
| 203831000000104 | Norwegian language interpreter needed |
| 203841000000108 | Pashto language interpreter needed |
| 203901000000102 | Polish language interpreter needed |
| 203911000000100 | Portuguese language interpreter needed |
| 203961000000103 | Panjabi language interpreter needed |
| 203971000000105 | Russian language interpreter needed |
| 203981000000107 | Serbian language interpreter needed |
| 203991000000109 | Shona language interpreter needed |
| 204011000000102 | Sinhala language interpreter needed |
| 204021000000108 | Somali language interpreter needed |
| 204031000000105 | Spanish language interpreter needed |
| 204041000000101 | Swahili language interpreter needed |
| 204051000000103 | Swedish language interpreter needed |
| 204081000000109 | Sylheti language interpreter needed |
| 204111000000101 | Tagalog language interpreter needed |
| 204131000000109 | Tamil language interpreter needed |
| 204151000000102 | Thai language interpreter needed |
| 204171000000106 | Tigrinya language interpreter needed |
| 204191000000105 | Turkish language interpreter needed |
| 204201000000107 | Ukrainian language interpreter needed |
| 204211000000109 | Urdu language interpreter needed |
| 204221000000103 | Vietnamese language interpreter needed |
| 204241000000105 | Yoruba language interpreter needed |
| 303601000000100 | Telephone interpreting service used |
| 314431000 | Interpreter present |
| 315593009 | Need for interpreter |
| 315594003 | Interpreter needed |
| 343711000000101 | Flemish language interpreter needed |
| 343771000000106 | Kutchi language interpreter needed |
| 352901000000108 | Slovak language interpreter needed |
| 352931000000102 | Ndebele language interpreter needed |
| 353881000000101 | Romanian language interpreter needed |
| 353921000000107 | Bulgarian language interpreter needed |
| 359641000000107 | Mongolian language interpreter needed |
| 359671000000101 | Moldavian language interpreter needed |
| 359701000000102 | Marathi language interpreter needed |
| 359731000000108 | Maltese language interpreter needed |
| 359761000000103 | Luganda language interpreter needed |
| 359791000000109 | Ethiopian language interpreter needed |
| 359821000000104 | Brawa language interpreter needed |
| 359851000000109 | Kirghiz language interpreter needed |
| 359881000000103 | Iban language interpreter needed |
| 359971000000108 | Macedonian language interpreter needed |
| 360011000000101 | Malagasy language interpreter needed |
| 360071000000106 | Latvian language interpreter needed |
| 360101000000102 | Kannada language interpreter needed |
| 360131000000108 | Kinyarwanda language interpreter needed |
| 360161000000103 | Malay language interpreter needed |
| 360191000000109 | Kashmiri language interpreter needed |
| 360221000000102 | Kazakh language interpreter needed |
| 360281000000101 | Javanese language interpreter needed |
| 360311000000103 | Inuktitut language interpreter needed |
| 360341000000102 | Interlingue language interpreter needed |
| 360371000000108 | Lao language interpreter needed |
| 360431000000104 | Hungarian language interpreter needed |
| 361441000000100 | Guarani language interpreter needed |
| 361471000000106 | Georgian language interpreter needed |
| 361501000000104 | Frisian language interpreter needed |
| 361791000000100 | Faroese language interpreter needed |
| 361821000000105 | Fijian language interpreter needed |
| 361881000000106 | Esperanto language interpreter needed |
| 361911000000106 | Estonian language interpreter needed |
| 361971000000101 | Danish language interpreter needed |
| 362041000000101 | Catalan language interpreter needed |
| 362071000000107 | Belarusian language interpreter needed |
| 362111000000101 | Indonesian language interpreter needed |
| 362141000000100 | Breton language interpreter needed |
| 362171000000106 | Bislama language interpreter needed |
| 362201000000107 | Bihari language interpreter needed |
| 362231000000101 | Assamese language interpreter needed |
| 362261000000106 | Armenian language interpreter needed |
| 362371000000109 | Burmese language interpreter needed |
| 362571000000102 | Afrikaans language interpreter needed |
| 362611000000106 | Azerbaijani language interpreter needed |
| 362641000000107 | Basque language interpreter needed |
| 362691000000102 | Afar language interpreter needed |
| 362721000000106 | Abkhazian language interpreter needed |
| 362821000000104 | Zulu language interpreter needed |
| 362961000000101 | Uzbek language interpreter needed |
| 362991000000107 | Oromo language interpreter needed |
| 363021000000102 | Yiddish language interpreter needed |
| 363061000000105 | Rundi language interpreter needed |
| 363141000000105 | Tibetan language interpreter needed |
| 363181000000102 | Tsonga language interpreter needed |
| 363301000000109 | Twi language interpreter needed |
| 363341000000107 | Telugu language interpreter needed |
| 363461000000100 | Tongan language interpreter needed |
| 363551000000104 | Turkmen language interpreter needed |
| 363591000000107 | Slovenian language interpreter needed |
| 363621000000105 | Swati language interpreter needed |
| 363651000000100 | Southern Sotho language interpreter needed |
| 363681000000106 | Tajik language interpreter needed |
| 363711000000105 | Sindhi language interpreter needed |
| 363781000000103 | Sundanese language interpreter needed |
| 363841000000104 | Samoan language interpreter needed |
| 363941000000108 | Tswana language interpreter needed |
| 364011000000105 | Quechua language interpreter needed |
| 364051000000109 | Sango language interpreter needed |
| 364141000000107 | Uighur language interpreter needed |
| 364211000000103 | Oriya language interpreter needed |
| 364251000000104 | Nepali language interpreter needed |
| 364311000000108 | Occitan language interpreter needed |
| 364351000000107 | Nauru language interpreter needed |
| 364481000000106 | Romansh language interpreter needed |
| 364511000000100 | Xhosa language interpreter needed |
| 423785008 | Provision of interpreter/translator services |
| 445075008 | Request for language interpreter service |
| 736790000 | Interpreter booked |
| 745664000 | Requires telephone language interpreter service |
| 787661000000108 | Interpreter booked |
| 945731000000104 | Hands-on signing interpreter needed |
| 972511000000109 | Aragonese interpreter needed |
| 972521000000103 | Avaric interpreter needed |
| 972531000000101 | Avestan interpreter needed |
| 972541000000105 | Bambara interpreter needed |
| 972551000000108 | Bashkir interpreter needed |
| 972561000000106 | Bosnian interpreter needed |
| 972571000000104 | Chamorro interpreter needed |
| 972581000000102 | Chechen interpreter needed |
| 972591000000100 | Chinese interpreter needed |
| 972601000000106 | Church Slavic interpreter needed |
| 972641000000109 | Dhivehi interpreter needed |
| 972651000000107 | Ewe interpreter needed |
| 972671000000103 | Fulah interpreter needed |
| 972691000000104 | Herero interpreter needed |
| 972711000000102 | Ido interpreter needed |
| 972721000000108 | Interlingua interpreter needed |
| 972731000000105 | Irish interpreter needed |
| 972751000000103 | Kikuyu interpreter needed |
| 972781000000109 | Kongo interpreter needed |
| 972801000000105 | Latin interpreter needed |
| 972831000000104 | Luxembourgish interpreter needed |
| 972851000000106 | Manx interpreter needed |
| 972881000000100 | Navajo interpreter needed |
| 972911000000100 | North Ndebele interpreter needed |
| 972931000000108 | Norwegian Bokmål interpreter needed |
| 972951000000101 | Nyanja interpreter needed |
| 973001000000108 | Ossetian interpreter needed |
| 973061000000107 | Serbo-Croatian interpreter needed |
| 973121000000100 | Venda interpreter needed |
| 973131000000103 | Volapük interpreter needed |
| 973141000000107 | Walloon interpreter needed |
| 973161000000108 | Wolof interpreter needed |
